# Persistence of Extended Spectrum β-Lactamase-Producing Enterobacterales in the Gut Microbiome of Healthy Newborns

**DOI:** 10.64898/2026.09.01.26361559

**Authors:** Weitao Shuai, Leena B. Mithal, Aspen Kremer, Abigail Aron, Alima Sajwani, Danielle Huntinghouse, Erica M. Hartmann, Mehreen Arshad

**Author notes:** shared first authorship (Weitao Shuai, Leena B. Mithal).

## Abstract

The global prevalence of Extended-spectrum β-lactamase-producing Enterobacterales (ESBL-E) colonization is increasing. However, it is unclear whether ESBL-E persist and if that is associated with an altered gut microbial ecology especially in early life where the developing microbiome may not provide the same colonization resistance as in adults. In this study, we collected longitudinal infant gut microbiome samples at delivery and in the nonclinical home setting in Chicago, Illinois, U.S.A, aiming to disentangle how genetic factors pertaining to the ESBL-E, as well as the surrounding gut ecology, influences persistence in the infant gut microbiome. We observed not only a higher-than-expected prevalence of ESBL-E in healthy infant gut microbiomes, but also a trend of ESBL-E persistence once colonized. Microbial communities showed higher dissimilarity between ESBL-E positive and negative infant gut microbiome at earlier time points. Although dissimilarity decreased over time, we present evidence that ESBL-E persist even when traditional detection methods are negative.

## 1. Introduction

The gut is a major reservoir of Gram-negative bacteria (GNB), such as *Escherichia coli*^1–3^, which serve as pathobionts, i.e., microorganisms that are part of the microbiota but have the potential to cause disease. However, these bacteria can persistently colonize the infant gut even in the absence of overt infections.

In women of reproductive age, *E. coli* and other GNB colonize the gut and occupy perianal, periurethral, and vaginal niches, and thus are proximal to the emerging newborn during vaginal birth^4^. Rates of maternal colonization with Extended-spectrum β-lactamase-producing Enterobacterales (ESBL-E) in high-income countries vary from 2.9% in Norway to 14% in the US^5–7^. In low-and middle-income countries (LMICs) ESBL-E colonization rates can be as high as 94% among peripartum mothers and 89% among newborns^8^. In a systematic review of studies in high- and upper-middle-income countries, the rate of perinatal transmission of multi-drug resistant GNB from colonized mothers to their infants was 27%^5^.

The maternal microbiota is a key determinant of the infant’s microbiome, especially in the first six months^9^. This period is critical for postnatal development, including of metabolic, immunologic, and neurocognitive systems^10^. An overabundance of pathobionts, such as *E. coli*, and a decrease in important commensals such as *Bifidobacteria* spp., *Lactobacillus* spp., and *Bacteroides* spp., can result in alterations of the gut microbiota (or dysbiosis) in infants and adverse health outcomes^11–13^.

Contemporary ESBL *E. coli* readily transmit and persistently colonize human hosts^14,15^. In animal models, these strains outcompete commensal *E. coli* in the gastrointestinal tract^15,16^. While global ESBL *E. coli* carriage in healthcare settings increased 3-fold from 7% in 2001–05 to 25.7% in 2016–20, in community settings it increased 10-fold from 2.6% to 26.4%^17^.

Detection of colonizing ESBL-E from clinical samples often relies on culture-based or PCR methods. However, the low proportion of culturable bacteria^18^ and PCR-target limitations constrain our ability to detect ESBL-E colonization. Therefore, ESBL-E prevalence in human gut microbiomes is likely underestimated. Development in sequencing technologies and bioinformatics enables characterization of the entire microbial community taxonomically and functionally. ESBL-E identification via metagenomic sequencing and assembly-based analysis is independent of their culturability but requires sufficient host-free biomass and sequencing depth. Here we sought to resolve ESBL-E colonization status in gut microbiome samples using both culture-based and metagenome-based methods, providing insights of current prevalence and characteristics of ESBL-E colonization.

Given the rapid increase in community acquired ESBL-E we have investigated a cohort of healthy mother-infant dyads with enough potentially ESBL-E colonized individuals in this study to allow reasonable comparisons between ESBL-E colonized *versus* non-ESBL-E colonized gut microbiome. We hypothesize that healthy neonates acquire ESBL-E which persist in their gut microbiome even without antibiotic exposure and that infant, maternal, and environmental factors, and pathogen-specific characteristics of ESBL-E collectively drive ESBL-E colonization and persistence in early life. We also hypothesize that ESBL-E colonization may drive distinct taxonomic and functional alterations in the gut microbiota, which could result in sustained ecological change and long-term health consequences. With the metagenome, serial isolates, and survey data collected from this cohort over eight months, we aim to (1) resolve ESBL-E colonization status, (2) demonstrate whether ESBL-E colonize and persist in healthy infant guts, (3) determine key factors that influence ESBL-E colonization and persistence, and (4) unveil microbial features of ESBL-E colonized infant gut microbiota.

Our results demonstrate the utility of metagenomic sequencing, including targeted read mapping, for longitudinal study of strain-level colonization. We also corroborate the general divergence and diversification of the gut microbiome over time, which may artefactually depress the ability to detect low-abundance ESBL-E.

## 2. Results

### 2.1 Overview of the cohort and microbiome

The cohort study recruited pregnant individuals and their newborn infants at Northwestern Medicine Prentice Women’s Hospital (PWH) in Illinois, USA (**Figure 1A)**. The inclusion of dyads was stratified by ceftriaxone-resistant (CefR) culture outcomes; 30 dyads had at least one CefR positive (CefR+) sample and the other 31 dyads were CefR negative (CefR-) for all samples collected for mothers and infants throughout the study.

**Figure 1.**
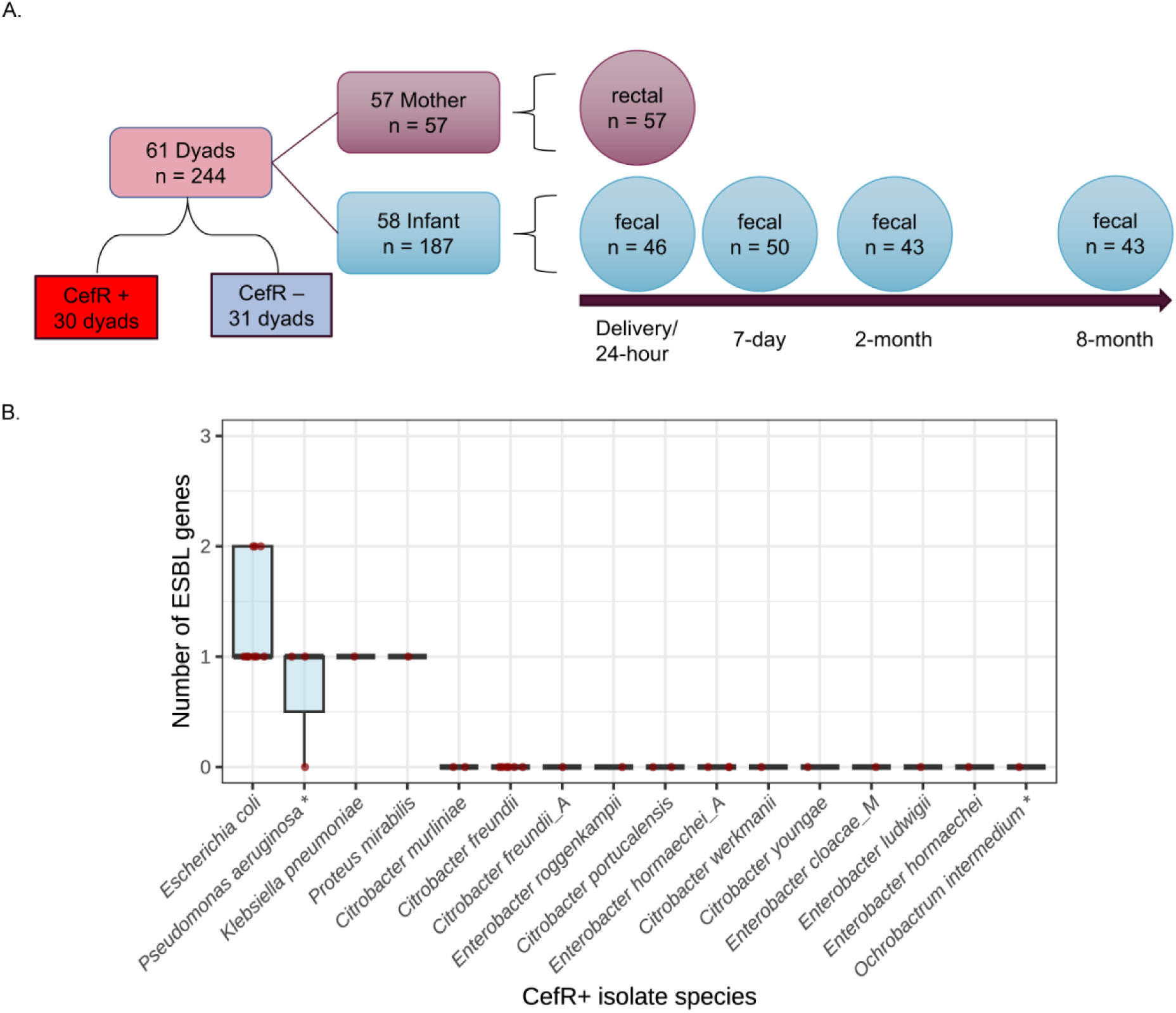
Overview of microbiome samples collected from the cohort and their culture-based results. (A) Mother-infant dyad stratification and number of samples at each time point. (B) Number of extended-spectrum β-lactamase (ESBL) genes carried by ceftriaxone-resistant (CefR+) isolates from all samples collected from the cohort. Species labeled with “*” (*Pseudomonas aeruginosa* and *Ochrobactrum intermedium*) are not Enterobacterales.

Although dyads included in this study were stratified by CefR status, not all CefR+ isolates proved to be ESBL-E (**Figure 1B**). A**Figure *1***mong the total 35 CefR+ isolates, *E. coli* was the most common species (9 isolates; **Figure 1B**). The majority of CefR+ isolates were Enterobacterales (88.6%) except for three *Pseudomonas aeruginosa* isolates and one *Ochrobactrum intermedium* isolate. Interestingly, only *E. coli*, *Klebsiella pnemoniae*, *Proteus mirabilis*, and some isolates of *P. aeruginosa* (non-Enterobacterales) harbored known ESBL genes, which consisted of 37.1% (13/35) of the total CefR+ isolates. Although many CefR+ Enterobacterales isolates do not harbor ESBL genes, they have at least one beta-lactamase gene (**Figure S1**).

Taxonomic profiling of all the metagenome samples resulted in a total of 1,100 species based on species-level genome bins (SGBs). The alpha diversity indexes showed significant difference between groups (ANOVA, *p* < 2.2×10^-16^), with increasing trends for infant samples over time and highest alpha diversities for mother samples (**Figure S2**).

### 2.2 ESBL-E colonization was observed in healthy infant gut microbiome mainly through metagenome-based identification

Stratification of the cohort based on CefR was insufficient for defining ESBL-E colonization. Therefore, we created a new indicator for putative ESBL-E colonization. The final ESBL-E status was resolved using three approaches: culture/WGS-based, assembly-based, and read mapping-based classification. Briefly, a sample is considered ESBL-E positive for culture/WGS-based if it resulted in one or more CefR+ isolate(s) and the isolate(s) were identified to be ESBL gene-harboring Enterobacterales. For metagenome samples where we were able to obtain scaffolds and high-quality metagenome-assembled genomes (MAGs), when taxonomy was assigned as Enterobacterales and an ESBL gene was identified from that scaffold or MAG, the sample was considered ESBL-E positive for assembly-based classification. In addition, clean short reads were mapped to ESBL-E assemblies for further ESBL-E identification, labeled as read mapping-based classification. On its own, the targeted read mapping strategy would likely result in a high number of false positives. Thus, we also included qPCR experiments for selected StudyIDs to verify our identification of ESBL genes from the metagenome samples. ESBL genes were identified even though no ESBL-E assemblies were resolved (**Figure S3**). Comparing the agreements between several ESBL-E classification methods (**Figure S4**), read mapping-based approach with permissive ESBL-E assembly coverage and depth and stringent ESBL gene coverage and depth had better agreement with qPCR validation. Therefore, this method was included for ESBL-E classification in addition to culture/WGS-based and assembly-based methods. Details for the comparisons of the approaches are included in **Supplementary Methods**.

A sample was considered positive if either culture/WGS-based, assembly-based, or read mapping-based classification was positive, and negative if all approaches identified no ESBL-E isolates, ESBL-E assemblies, or low coverage and depth of ESBL-E. Details for these approaches are described in **Methods and Materials**. Downstream analyses of the cohort’s gut microbiome are based on this final ESBL-E status classification.

All maternal ESBL-E colonization (5 out of 57 mothers) was identified through the culture/WGS-based approach, while most infant ESBL-E colonization was identified through the metagenome-based approaches (assembly-based and read mapping-based). Among the 111 ESBL-E positive infant microbiomes, only 5 had ESBL-E isolates identified through culture/WGS (**Figure 2A**). Among the 54 dyads with a maternal sample and at least one infant sample, 43 had at least one ESBL-E positive maternal or infant gut microbiome sample (**Figure 2B**), which is much higher compared to the culture-based CefR screening alone.

**Figure 2.**
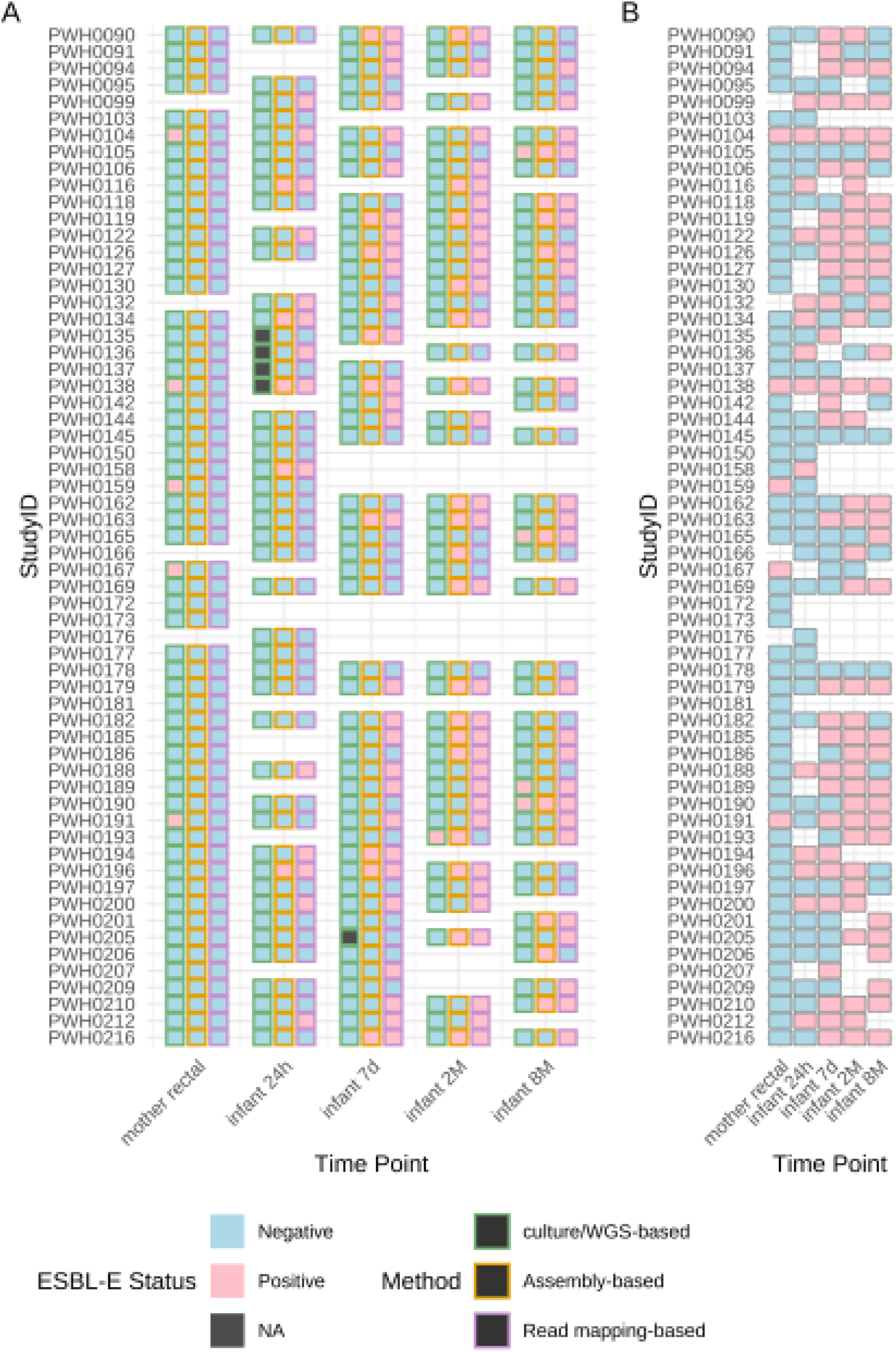
Identification of ESBL-E in the microbiome samples of mother-infant dyads using culture/WGS-based, assembly-based, and read mapping-based methods (A) and the final ESBL-E status for each sample (B). Black boxes indicate that culture results are not available. Empty blocks indicate no samples were collected at the time point. For each sample, if either identification method is positive in panel A, the final ESBL-E status in panel B is considered positive.

### 2.3 ESBL-E are persistent in healthy infant gut microbiomes

Among the 5 ESBL-E positive mothers, 3 had ESBL-E positive infants (StudyID PWH0104, PWH0138, and PWH0191). This small number of ESBL-E positive mother-infant dyads limited our ability to investigate perinatal transmission. However, this result is consistent with our hypothesis that pathways other than perinatal transmission contribute to ESBL-E colonization during early infancy.

Here we focus on ESBL-E colonization and persistence in healthy infant gut microbiomes. There were 20 infants with at least 3 ESBL-E positive time points (**Figure 2B**). By mapping metagenome reads to the ESBL-E assemblies, we obtained the coverage and truncated average depth (TAD) of the ESBL-E assembly as well as the coverage and TAD of the ESBL gene locus (details in **Methods and Materials** and **Supplementary Methods**). Higher coverage and TAD correspond to higher completeness and abundance in the metagenome sample, respectively. By investigating all available metagenome samples of these dyads for ESBL-E colonization based on short read mapping results, we observed the same ESBL-E typing appearing in the same individual for 14 infants over time (**Figure 3A**). Tracking the relative abundance of the ESBL-E assemblies over time for these infants, higher abundance was observed at earlier time points compared to later time points (**Figure 3B**).

**Figure 3.**
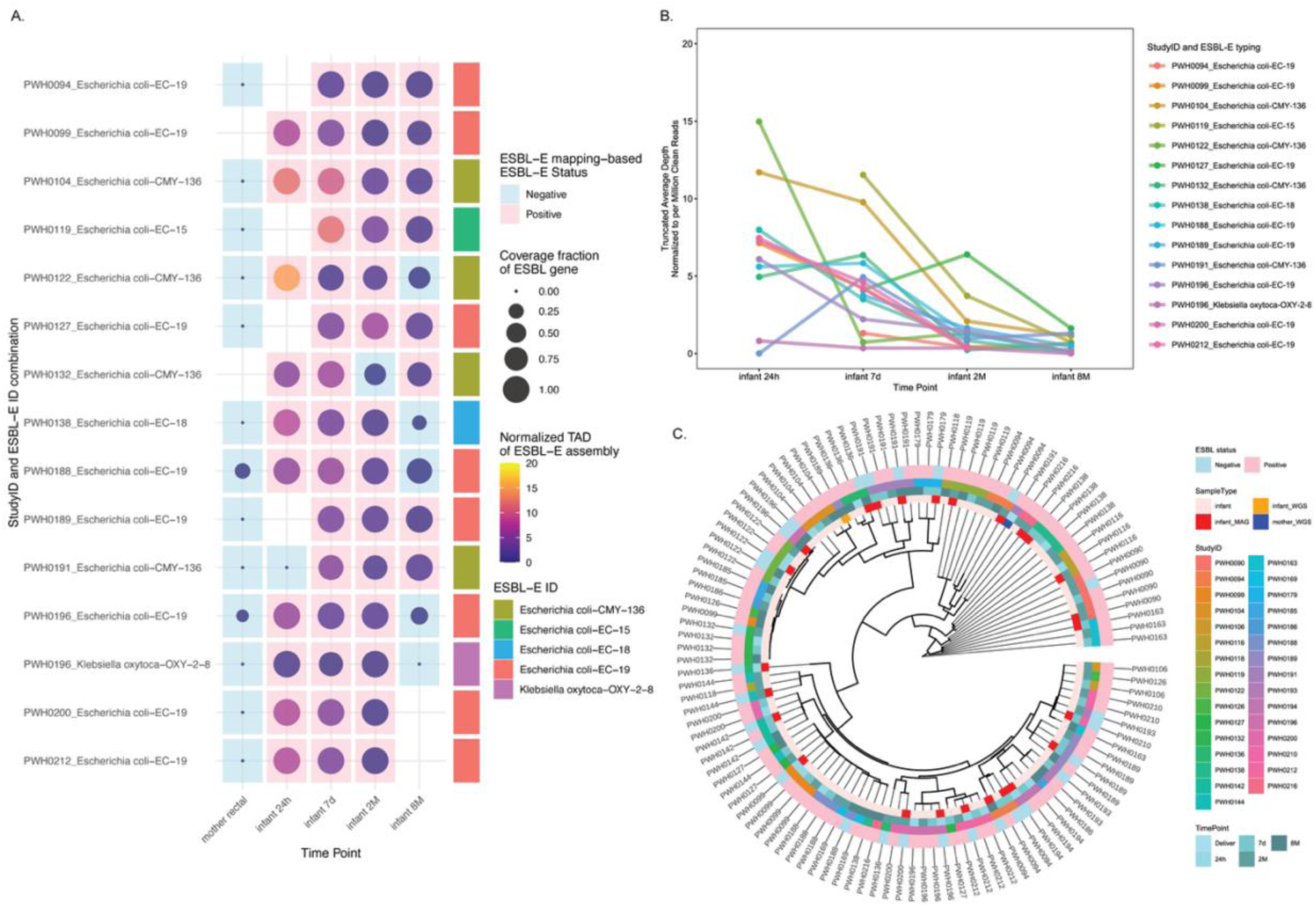
Patterns of ESBL-E persistence in infant gut microbiome. Mother-infant dyads were numbered by StudyID. (A) Metagenome mapping results for individuals with at least three ESBL-E positive infant time points. The color of the background indicates ESBL-E status from read mapping-based approach. The ESBL-E typing was colored for each StudyID. Truncated average depth (TAD) normalized to per million clean reads of the ESBL-E assemblies and ESBL gene coverage fraction are indicated by the circle color and size, respectively. The empty blocks indicate no samples were collected for the time point. (B) Normalized TAD of ESBL-E assemblies over time for the infant individuals presented in panel A. (C) Phylogenetic tree constructed from metagenomic short reads using metagenome assembled genomes (MAG) or ESBL-carrying isolate genomes (WGS) of *E. coli* from the cohort as reference genomes. Phylogeny was determined based on alignments of *E. coli* marker genes.

We further resolved ESBL-E persistence status for each infant individual by investigating each sample’s ESBL-E status and taxon-ESBL family typing from metagenome-based results (**Table S1; Table 1**)**Table *1***. *E. coli* with EC family ESBL and *Klebsiella* spp. with OXY family ESBL co-existed in 11 infants of this cohort, with *E. coli*-EC presented much higher persistence compared to *Klebsiella* spp.-OXY (6 infants versus 1 infants).

**Table 1.** Persistence status of infant individuals for different Enterobacterales with extended-spectrum beta-lactamase gene families.

| ESBL-E | Persistent | Transient |
| --- | --- | --- |
| *Escherichia coli*-EC family ESBL | 20 | 17 |
| *Escherichia coli*-CMY family ESBL | 3 | 9 |
| *Klebsiella* spp.-OXY family ESBL | 3 | 11 |
| *Citrobacter freundii*-CMY family ESBL | 1 | 3 |
| *Escherichia coli*-CTX-M family ESBL | 0 | 2 |

ESBL-*E. coli* was the most prevalent ESBL-E that persisted in the cohort. The strain-level population structure showed that *E. coli* strains from the same individual tend to have smaller phylogenetic distances, clustering together in the *E. coli* phylogenetic tree (**Figure 3C**). Normalized pairwise phylogenetic distance further provided evidence of stable colonization with a single strain per individual, rather than sporadic acquisition of different strains over time (**Figure S5**). This clustering of strains from same individual is more prominent compared to the clustering by ESBL status (indicated by the outer ring of the phylogenetic tree in **Figure 3C**).

### 2.4 Association of clinical and demographic variables with ESBL-E colonization and persistence among infants

We investigated the association between demographic and clinical variables and infant ESBL-E colonization at early (first 7 days of life), 2 months, and 8 months (**Table 2**). Race and Latine ethnicity were not associated with ESBL-E colonization at any timepoint. Maternal health factors including gestational DM, antibiotics during pregnancy, probiotic use, acid-blocking medication, and birth outside the U.S. use were not significantly associated with infant ESBL-E colonization. Infant sex and probiotic use at any timepoint were not associated with ESBL-E colonization. While our cohort skewed toward high proportion of vaginal delivery, having a vaginal delivery was associated with neonatal ESBL-E colonization within the first week of life (*p* = 0.03) and ESBL-E presence at any time point, driven by this early finding (*p* < 0.001). Formula only feeding in the first 2 months of life was associated with ESBL-E colonization at 8 months of age (*p* = 0.043). Breastmilk versus formula feeding at 8 months of life did not have the same association, noting that by 8 months infants are eating solids, but showed a trend toward association with persistence of ESBL-E strain. Infants who received only formula had higher rate of transient ESBL-E colonization; those who received breastmilk only had more persistent ESBL-E strains (*p* = 0.054).

**Table 2.**
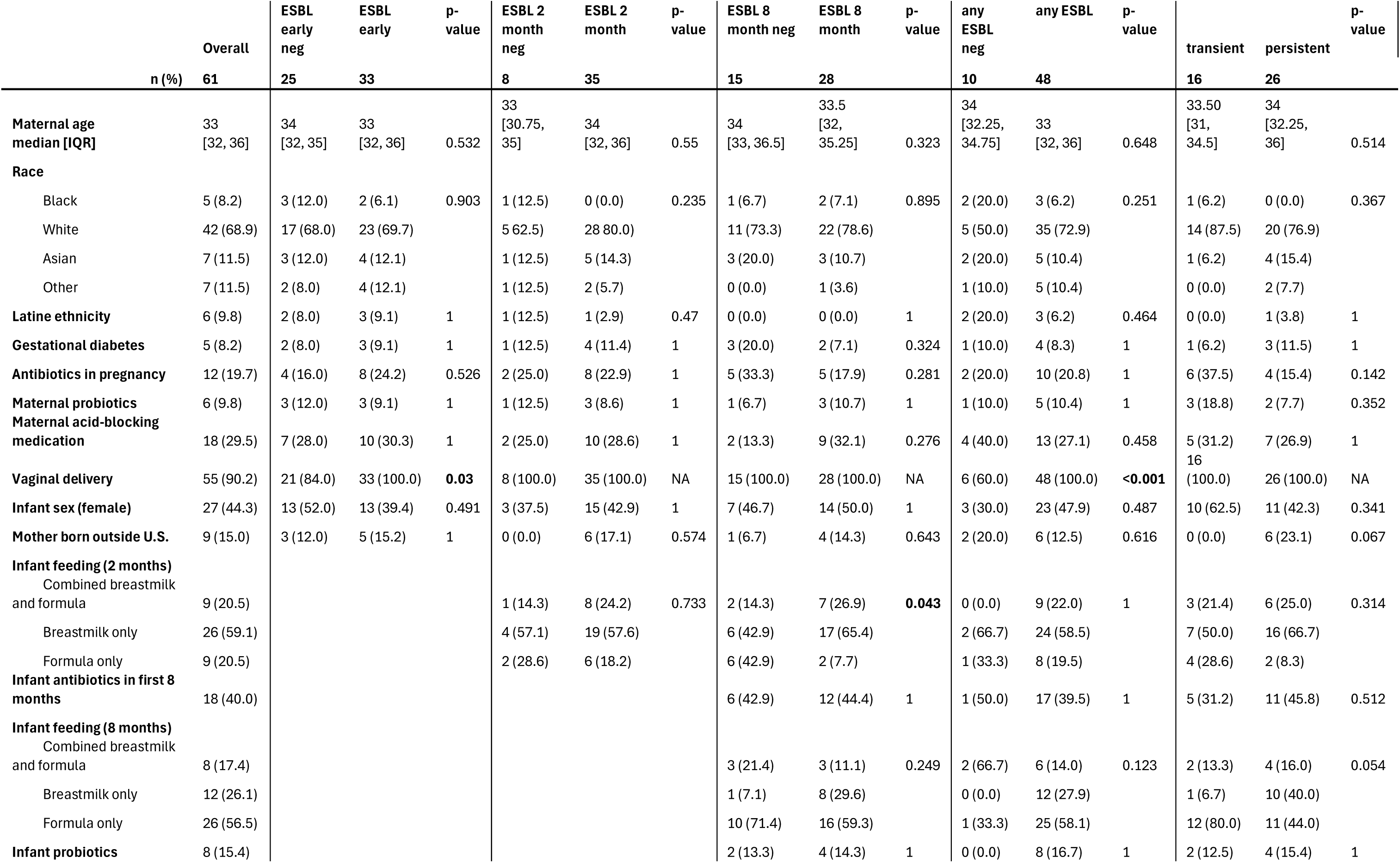
Infant ESBL-E colonization status by demographic and clinical variables.

### 2.5 Taxonomy of the ESBL-E and co-colonizing organisms affect colonization and persistence

Although the ESBL gene alone may not be the defining feature at the strain level, we hypothesize that some genetic elements in the genomes such as mobile genetic elements (MGEs) and virulence factors (VFs) could contribute to maintaining ESBL genes in the strains that colonized healthy infant gut microbiome. Different types of ESBL genes tended to co-exist with different VFs in ESBL-E genomes with a species-specific pattern (**Figure 4A**). Bla-EC, bla-CTX, and bla-CMY ESBL genes were most frequently identified in *E. coli* genomes and bla-OXY ESBL genes were identified only in *K. oxytoca*, *K. michiganensis*, and *K. grimontii* genomes, which aligns with the grouping shown in the network (**Figure S6A**). The presence/absence pattern of VFs showed species as a stronger differentiator compared to ESBL types (**Figure S7**). Among all the Enterobacterales genomes recovered, the associations between ESBL gene and VF gene co-occurrence are not statistically significant (**Figure S8)** except for bla-OXY-1-1 and VF gene Pla (phi coefficient, Fisher’s exact test *p* = 0.016). ESBL genes of the same beta-lactamase gene families showed similarities in co-occurrence pattern with VF genes, but not with MGEs (**Figure S6**).

**Figure 4.**
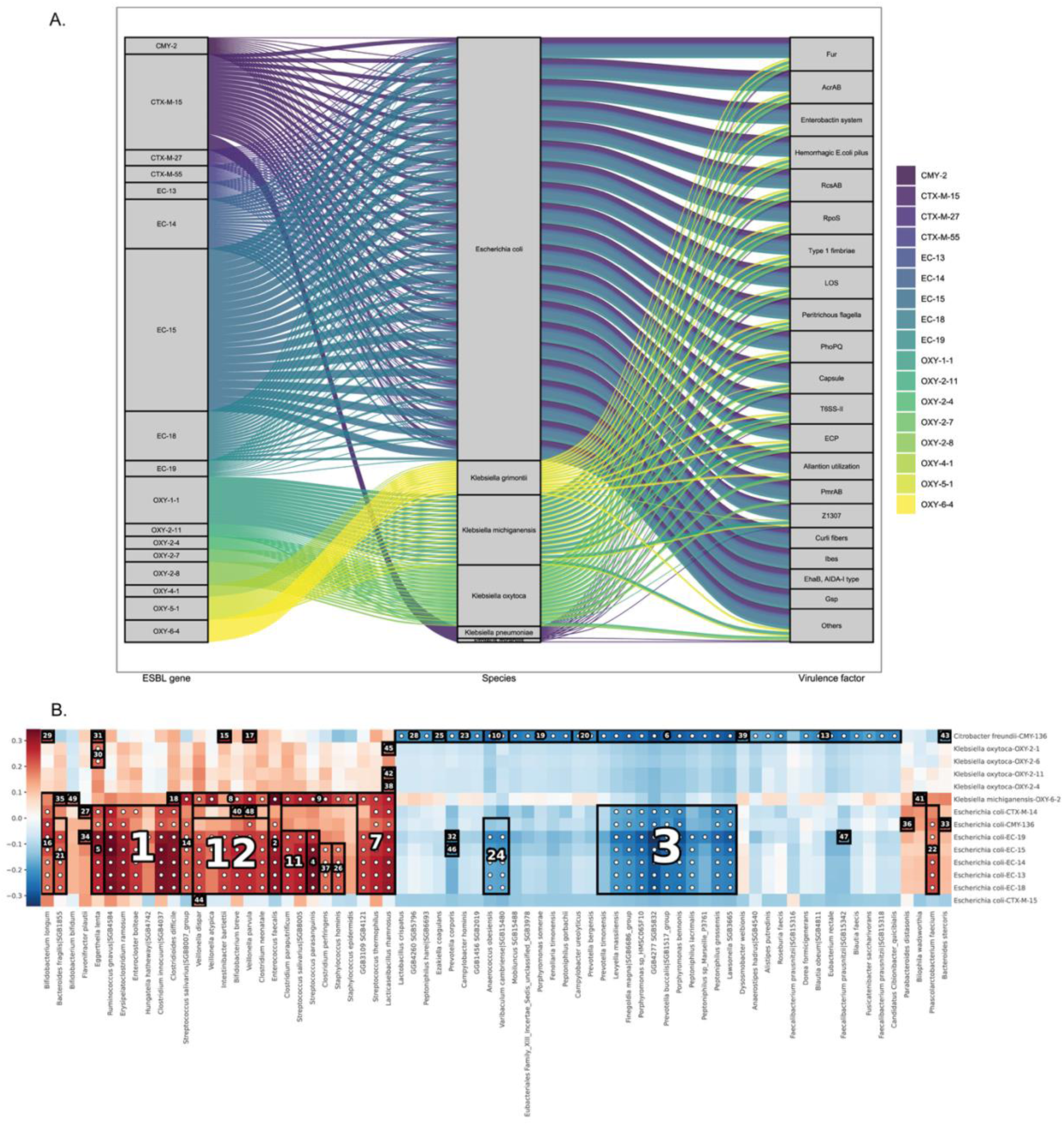
Genomic context and ecological associations of ESBL-E. (A) Co-existence of ESBL genes and VFs in the same MAGs. Each band represents an instance where an ESBL gene and a VF are present in the same genome. The middle column represents Enterobacterales genomes carrying both ESBL genes and VFs with species classification. (B) Hierarchical all-against-all associations between ESBL-E assemblies and taxa at species-level genome bins with more than 10% prevalence in the cohort gut microbiome.

A total of 1051 MAGs and 35 CefR+ isolate genomes (culture-WGS generated) were generated from the cohort metagenomes, of which 165 were Enterobacterales. While 29 Enterobacterales species were identified, only 6 species harbored ESBL genes (**Figure S9**), which comprised 25.5% of the Enterobacterales genomes. The most prevalent ESBL-E was *E. coli*, followed by *K. michiganensis* and *K. oxytoca*. While ESBL genes were not identified in most *E. coli* genomes, they were detected in all genomes of *K. michiganensis* and *K. oxytoca* (**Figure S9**). Similar to strain-level population structure, gene family profiles of *E. coli* ESBL positive or negative samples did not show evident clustering (**Figure S10**). Gene profiles of *K. michiganensis* and *K. oxycota* showed a higher level of strain-strain similarities to reference genomes, with some clustering of ESBL positive or negative samples (**Figure S11**).

The top-ranked positive association cluster from hierarchical all-against-all statistical analysis (**Figure 4B**) was between ESBL-carrying *E. coli* assemblies and a group of microbes including *Ruminococcus gnavus* (SGB4584), *Erysipelatoclostridium ramosum*, *Clostridium innocuum* (SGB4037), *Clostridioides difficile*, *Enterocloster bolteae*, and *Hungatella hathewayi* (SGB4742). The top ranked negative association cluster was between ESBL-carrying *E. coli* assemblies and a group including species of *Porphyromonas*, *Peptoniphilus*, *Prevotella*, *Levyella*, *Finegoldia*, and *Lawsonella*.

### 2.6 Distinct features of infant gut microbiome colonized with ESBL-Es

To investigate whether ESBL-E colonization would affect the gut microbial community, alpha- and beta-diversities of the mother and infant samples were compared between ESBL-E status. No significant difference was observed for mother or infant gut microbial richness at the species level (Wilcoxon rank-sum test *p* > 0.1, labeled in **Figure S12**). Permutational Multivariate Analysis of Variance (PERMANOVA) of all mother and infant samples showed that the gut microbiome ESBL-E status is a weak factor explaining microbial community dissimilarity (**Figure S13A**; R^2^ = 0.038, *p* = 0.0001). No statistically significant difference between ESBL-E status was observed for mother samples (**Figure S13B**, R^2^ = 0.016, *p* = 0.66). ESBL-E status (R^2^ = 0.058 and *p* = 0.0001) explains less variance than time (R^2^ = 0.090, *p* = 0.0001) for infant gut microbiomes. Microbial community dissimilarity between ESBL-E positive *versus* ESBL-E negative infant gut microbiome showed a decreasing trend over time (**Figure 5**). Kruskal-Wallis test for Bray-Curtis dissimilarities between different time point groups (*p* < 0.001) and pairwise Wilcox test showed statistically significant differences between any two time points (*p* values < 0.001, **Table S2**).

**Figure 5.**
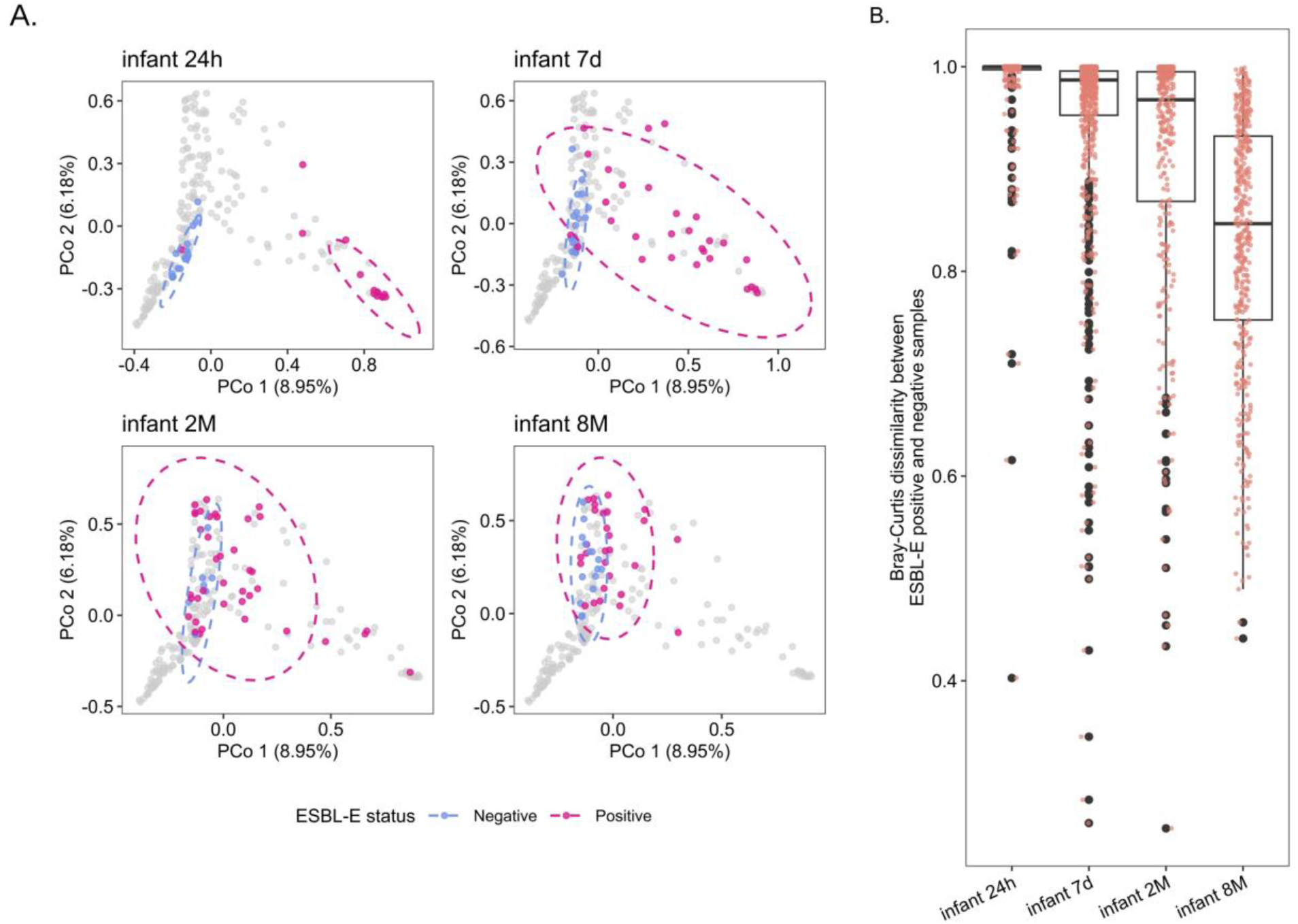
Bray-Curtis dissimilarity between infant gut microbiome metagenome samples. (A) Principal coordinate analysis (PCoA) of infant gut microbial communities grouped by time points and labeled with their ESBL-E status. (B) Dissimilarities between ESBL-E positive *versus* ESBL-E negative infant gut microbial communities over time. Kruskal-Wallis test *p* < 2.2×10^-16^.

ESBL-E positive infant gut microbiomes showed higher relative abundance of Enterobacterales (**Figure S14A**). However, this may be an artefact of only being able to identify ESBL genes from those Enterobacterales present in the metagenomes with high enough relative abundance, which results in sufficient coverage and depth for mapping-based ESBL-E classification. With MaAsLin3 (Microbiome Multivariable Associations with Linear Models), we accounted for total clean read numbers from each metagenome sample as well as the repeated sampling of an individual and investigated key features of the infant gut metagenome with different ESBL-E status and time points. **Figure 6**ESBL-E positive infant gut microbiomes feature higher abundance and prevalence of *E. coli*, while *E. coli* abundance decreases over time (**Figure 6A, Figure S14BC**). *Enterococcus faecalis* has relatively stable prevalence in ESBL-E positive/negative infant samples and over time, with a decreasing trend in abundance over time, significant only in the 8-month gut microbiome (**Figure 6A, Figure S14D**). *B. longum*, a commensal gastrointestinal tract inhabitant, increases in prevalence over time (**Figure 6A, Figure S14E**).

**Figure 6.**
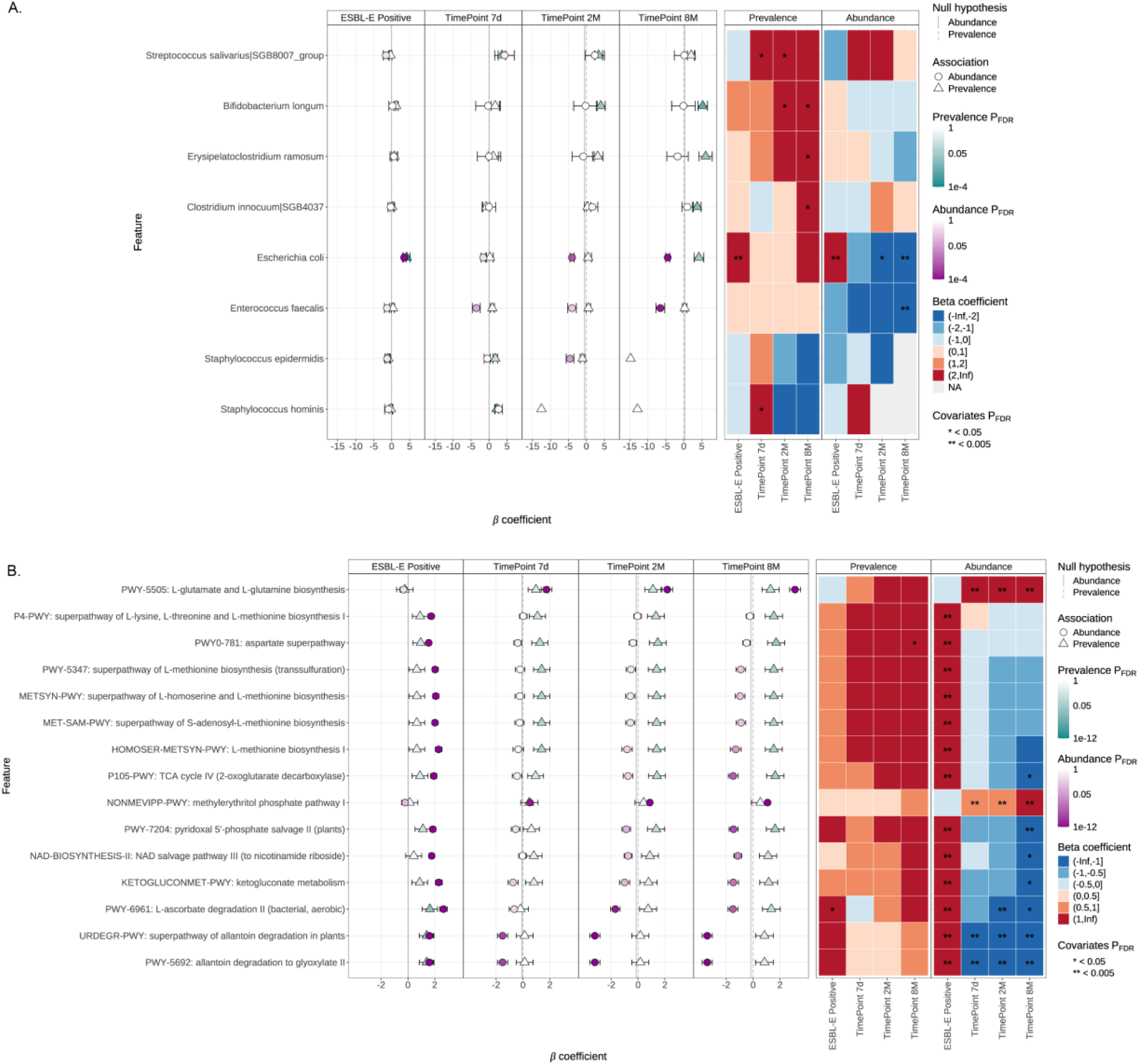
Microbial abundance and prevalence associations (A) and pathway abundance and prevalence associations (B) for infant gut microbiome of different time points and ESBL-E status. Reference level for time points is 24h and reference level for ESBL-E status is negative. Taxa (species-level genome bins) with significant associations and pathways of top 15 significant associations are shown as features in the summary plot. Metadata variables are ordered by their significant associations. Statistical significance metrics for the features are listed in **Table S3**.

Looking at the functional pathways identified in the infant gut metagenome samples, ESBL-E positive microbiome features elevated abundance in pathways for amino acids aspartate, L-lysine, L-threonine, L-methionine, and L-homoserine biosynthesis, TCA cycle IV, pyridoxal 5’-phosphate and NAD salvage, ketogluconate metabolism, and L-ascorbate (vitamin C) and allantoin degradation. Interestingly, the abundances of pathways showed the opposite enrichment/reduction pattern for the ESBL-E positive versus negative comparison and the later versus the first time point comparisons.

## 3. Discussion

### 3.1 Challenges in identifying ESBL-E colonization in gut microbiome of healthy individuals

The gut microbiome is recognized for its critical role in many developmental outcomes. However, it is still unclear where specific strains are acquired from and why only some persist. Paradoxically, this diversity is one reason it can be so difficult to study the gut microbiome at the strain level.

Of all taxa in the developing human gut, ESBL-E are of particular interest because of their resistance to antibiotics and potential to cause disease. The standard clinical approach using selective media is prone to both false positives, as demonstrated by the substantial discrepancies we observed between CefR+ culture results and whole genome sequencing-based verification, and false negatives, as demonstrated by our metagenomic sequencing data. The identification of phenotypically CefR+ isolates that do not carry ESBL genes indicates that mechanisms other than ESBLs also contribute to ceftriaxone resistance. Using short read-based taxonomic and functional profiling of metagenomes is limited in associating ESBL genes with their hosts. Therefore, assembly is needed to resolve this relationship. From MAGs or even some scaffolds, taxonomy can be assigned along with complete alignment of ESBL genes, allowing for ESBL-E identification. However, whether a high-quality assembly can be generated from a sample is affected by factors such as low clean read counts (low starting biomass such as seen in stools from newborns) and insufficient sequencing effort due to high diversity of the microbiome. It is thus not surprising that mapping short reads to the ESBL-E assemblies resulted in observable coverage and depth even from samples that yielded no ESBL-E assemblies. In addition, our ability to detect ESBL-E with targeted mapping would still be limited by the metagenome diversity and sequencing efforts. As the infant gut microbiomes develop and diversify, relative abundance of ESBL-E may drop to a point that sequencing data became insufficient to meet the threshold of detection. Therefore, the negativity of ESBL-E in later timepoints could be partially artefactual. A previous study on healthy infants showed a median clearance of ESBL-E colonization at 7.5 months^20^, which could be underestimated due to the limit of detection of culture-based methods as well.

With the mapping results, we found co-existence of ESBL-*E. coli* and ESBL-*Klebsiella* in some individuals, which was not detected by culture-based results. Co-colonization of ESBL-*E. coli* and ESBL-*Klebsiella* has been reported previously^21,22^, but their interactions and dynamics in the host microbiome were not investigated.

The nature of the maternal rectal samples at delivery and infant 24h samples resulted in lower-than-expected proportion of microbial biomass in these samples, hindering ESBL-E detection by a single method. This inconsistency of gut microbiome sample quality was also reflected in the total clean reads of metagenomes (**Figure S15**). ESBL-E classification across the three selected methods (**Figure 2A**) demonstrated that these different methods are complementary. While selective culture-based methods may work better in detecting ESBL-E for samples with low non-host biomass, metagenome-based methods have stronger ability in detecting ESBL-E that do not show up in selective culture. Integrating these different methods enables more comprehensive detection of ESBL-E from samples with various characteristics.

### 3.2 Implications of ESBL-E colonization in infant gut microbiome

We have not only demonstrated ESBL-E persistence in healthy infant gut microbiome after colonization, but also early ESBL-E detection within 48 hours of birth in most of the ESBL-E persistent individuals, which strongly suggests perinatal transmission. Our results demonstrated significant differences between the microbiomes of ESBL-E positive and negative healthy infants in their early lives. ESBL-E colonization drives collective functional features of the microbiome towards a different direction during development and affects the healthy infant gut microbial community composition overall. However, taxonomic richness remains similar, and the effect is more prominent in early time points, which indicates that early intervention for ESBL-E transmission may be more efficient in preventing adverse health outcomes. It is possible that as the infant gut microbiome develops and diversifies (**Figure S2**) the importance of ESBL-E colonization is attenuated by the colonization of other members of the gut microbiome.

Our clinical data analysis demonstrates that vaginal delivery is associated with early infant ESBL-E colonization compared to caesarean section. Our dataset had majority vaginal deliveries, and our capability to assess the durability of this difference was therefore limited. Therefore, our microbiota analyses at these timepoints include only infants delivered vaginally. Breastmilk feeding is associated with ESBL-E colonization and persistence, compared to formula only feeding. This may reflect continued exposure to maternal flora through breastfeeding. Breastmilk has several potential benefits including reduced infection, lower risk of certain chronic diseases, and improved neurodevelopmental outcomes^23–25^. Breastfeeding may also have a positive impact on gut microbiota, noted in both preterm and term infants^26–29^. Thus, while noting this association and potential route of ESBL-E colonization and persistence with breastmilk feeding, further investigation is required to understand this finding in a broader context.

Several studies of hospitalized patient populations have shown that ESBL-E positivity in the gut is associated with an altered microbiome^30^. It has been suggested that this is due to multiple and prolonged exposure to antibiotics and the selection pressure exerted by the hospital environment. However, studies in healthy adults in the community show that microbiome composition is not significantly different in individuals carrying ESBL-E compared to those that are negative^31^. Our results, therefore, suggest that the rapidly developing early infant microbiome is a unique environment where acquisition of ESBL-E is associated with significant changes in composition of the microbiota.

Among the significant results of differential prevalence and abundance for taxa in the infant gut microbiome, *B. longum* increased prevalence coincides with *E. coli* decreased abundance over time. *B. longum* is known to colonize infant gut microbiome in early life with high abundance, and strains of *B. longum* have been developed as probiotics due to their beneficial health effects.^32^ It is possible that colonization of *B. longum* exerts a protective effect from potential pathogens. However, positive associations between *B. longum* and ESBL-E abundances (**Figure 4B**, association cluster number 16) indicate that higher abundance of this probiotic bacterium does not necessarily outcompete ESBL-E. The co-colonization pattern of ESBL-E with *E. faecalis* has been described previously. In vitro and in vivo studies show that *E. faecalis* promotes the growth and survival of *E. coli* through the production of l-ornithine, the precursor needed to synthesize enterobactin (an iron-scavenging siderophore) and the formation of biofilms^33^. In infants this is especially concerning because both *E. coli* and *E. faecalis* are a major cause of invasive infections^34^.

Some of the species in the top-ranking positive association cluster with ESBL-E abundance are potentially pathogenic. *R. gnavus*, a prevalent member of the infant and adult microbiota, has been associated with Crohn’s disease^28^, inflammatory bowel diseases, and some other gut- and non-gut-related diseases^29^. *E. ramosum* (previously *Clostridium ramosum*) is usually a commensal microorganism in GI tract but can also be a pathogen. *C. innocuum* is part of the normal flora of GI tract but can also be an opportunistic bacterium^30^. *C. difficile* is known to cause serious diarrheal infections. *E. bolteae* (previously *Clostridium bolteae*) was found to be enriched in gut dysbiosis^31^. *H. hathewayi* (previously *Clostridium hathewayi*^32^) can cause rare infections. Among the species in the top-ranking negative association cluster, *Porphyromonas* are commonly found in human digestive tract^33^ and can be potential pathobionts^34^. *Peptoniphilus* is a part of the gut and vaginal microbiota. Prevotella are also commonly found in multiple human body sites including gut and vagina and were enriched in non-westernized populations^35^. *L. massiliensis* was found to have symbiotic relationships with *Veillonella parvula* in the intestinal microbiota of acute leukemia outpatients^36^. *F. magna* is part of the human GI microbiota^37^. *Lawsonella* is part of the normal human microbiota but also found to be associated with abscesses^38^.

### 3.3 Limitations

Establishing statistical significance of results in this study was hampered in part by the small cohort size and varying sample quality. Specifically, the metagenomic sequencing yielded high host cell contamination, resulting in low clean read yield for some of the maternal rectal samples and infant samples taken within 24 hours of birth. Bacterial biomass proportion was affected by inevitable interference such as the presence of ultrasound gel in mother rectal samples due to necessary procedure at the time of delivery and the fact that meconium instead of stool was produced within 24 hours of birth by newborns. Except for maternal and infant 24-hour samples, other infant time points had comparable clean sequence numbers for downstream analysis (**Figure S15**). Because gut microbiome diversity increases with time, but samples were sequenced in a single batch, the sequencing depth as indicated by coverage necessarily decreases with time. Overcoming this challenge requires an advanced estimate of gut microbiome diversity, as well as methods to avoid batch bias.

We also noticed a lack of plasmid-associated ESBL genes in metagenomic sequencing-based identification (**Figure S16**), indicating potential underrepresentation of plasmid-associated ESBL genes due to biased extraction, amplification, or assembly (plasmid contigs unable to assemble or to be binned in genomes) in the metagenome-based analysis pipeline. This further validated our approach to include both culture-WGS generated genomes and MAGs. Sample handling and processing should be carefully considered to preserve these specific features within the microbiome in future studies. These caveats should be addressed by experimental design and planning for sample collection and processing in future studies.

It was not feasible to identify the sources of non-perinatally transmitted ESBL-E without environmental and gut microbiome samples from other family members from this cohort. We propose to include such samples for future studies to better understand ESBL-E transmission pathways based on the findings from this study.

## 4. Methods and Materials

### Participant enrollment and cohort inclusion

This cohort was part of a prospective, observational study of pregnant women and microbial colonization of mother and their infants at Northwestern Medicine Prentice Women’s Hospital^46^. Exclusion criteria included anticipated preterm delivery (<35 weeks), fever during labor, scheduled cesarean delivery, and antibiotic use during labor before enrollment. Infants who were admitted to the neonatal intensive care unit were also excluded from subsequent prospective participation. The rationale for exclusion criteria was that antibiotic exposure during labor and neonatal intensive care hospitalization are known risk factors for drug resistant bacterial colonization, and our objective was to characterize a healthy cohort of infants in the community. Details on the participant enrollment, screening, and consent process during hospitalization are detailed in our prior manuscript.

### Sample processing

Rectal swabs were collected by the obstetric care providers during a routine exam before delivery. Infant stool samples were retrieved from diapers. The first sample (24h) was between 24-48 hours of life, before hospital discharge. Diapers at day 7-10 (7d), 2 months (2M), and 8 months (8M) were retrieved via courier from the patients’ homes. Details on ceftriaxone resistant isolates culturing and selection, whole genome sequencing sample processing, and shotgun metagenomic sequencing sample processing are described in **Supplementary Methods**.

While in the hospital, a questionnaire was administered including maternal medical conditions, country of origin, medications, and antibiotic receipt during pregnancy. Additional clinical data were manually abstracted from electronic medical records including delivery characteristics and infant nutrition. At the time of 2- and 8-month diaper collection, an online questionnaire was completed by participants via a secure REDCap online platform. This study was approved by the Institutional Review Board of Northwestern University STU2020331 before its initiation and conducted in accordance with the Declaration of Helsinki.

### Analyses for whole genome sequencing and metagenome-assembled assemblies

Assembly (WGS genome, MAG, or scaffold) taxonomic classification and functional annotation are described in **Supplementary Methods**. A list of extended-spectrum beta-lactamase (ESBL) genes from the National Center for Biotechnology Information (NCBI) Reference Gene Catalog (database version 2025-03-25.1) was used to identify ESBL genes from RGI outputs. An assembly is identified as an ESBL-E assembly if it carries an ESBL gene and is classified as Enterobacterales.

An ESBL-E assembly database was constructed from metagenome-generated assemblies as follows for downstream ESBL-E status identification for each gut metagenome sample. All ESBL-E MAGs were dereplicated at 99.5% identity to keep different ESBL gene typing using dRep^47^ v3.5.0. ESBL-E scaffolds that were not recruited to form ESBL-E MAGs in metaWRAP pipeline were also included in the ESBL-E assembly database. Bowtie2^48,49^ v2.5.4 was used to construct the database for mapping. Clean short reads of each metagenome sample were then mapped to this database with the alignment with the best MAPQ score reported. Mapping results were processed using SAMtools^50^ v1.14 and custom python scripts to obtain truncated average depth (TAD) and breadth of coverage for each ESBL-E assembly. Coverage of ESBL genes in the ESBL-E assemblies was obtained using BEDTools^51^ v2.31.1 with ESBL gene annotation information from RGI outputs. These mapping results were then used for ESBL-E status classification.

To investigate genomic context for ESBL genes in Enterobacterales, ESBL-E assemblies were first annotated using Prokka^52^ v1.14.6. Virulence factor (VF) protein sequences of full dataset were downloaded from the virulence factor database (VFDB) and used for DIAMOND^53,54^ (v0.9.22) database construction. Open reading frame amino acid sequences from Prokka were then searched against VFDB for VF identification using DIAMOND blastp using the following arguments: --id 80 --query-cover 80 --subject-cover 80 -e 0.00001 --strand both -k 1 --sensitive. MOB-suite^55^ v3.1.9 and MobileElementFinder^62^ v1.1.2 were used to identify mobile genetic elements from the ESBL-E assemblies. Custom R scripts were used to process the results.

### ESBL-E status classification

For each mother or infant microbiome sample, there are three methods to identify whether it is colonized with extended-spectrum beta-lactamase (ESBL)-producing Enterobacterales: (1) Culture/WGS-based ESBL-E status, (2) ESBL-E assembly presence-based ESBL-E status, and (3) ESBL-E assembly mapping-based ESBL-E status. For culture/WGS-based method, a sample is classified as ESBL-E positive if a CefR + isolate was obtained from this sample and the isolate’s genome was classified as Enterobacterales with ESBL gene(s) identified in the genome. For ESBL-E assembly presence-based method, a sample is classified as ESBL-E positive if a scaffold or MAG that was classified as Enterobacterales with ESBL gene(s) identified in the assembly sequence was assembled from this sample. For the ESBL-E assembly mapping-based method, a sample is classified as ESBL-E positive if the results of short reads mapping to the ESBL-E assembly database satisfy the following criteria: (1) the truncated average depth of a ESBL-E assembly normalized to per million clean reads (TAD_norm) > 0.2, (2) the breadth of coverage (range 0-1) of this ESBL-E assembly > 0.25, and (3) the coverage fraction (range 0-1) of the ESBL gene in this assembly > 0.8. Method validation can be found in Supplementary Methods. The final ESBL-E status for each sample was then determined based on the results of all three methods. A sample was classified as ESBL-E positive (+) if any one of the three methods yielded positive result; otherwise, it was classified as ESBL-E negative (-).

### ESBL-E persistence status classification

Persistence status was resolved for each infant by investigating metagenome-based ESBL-E results including taxon-ESBL typing and mapping outputs. Infant 24h and 7d samples were collapsed into one “within 7d” time point for each individual. If an ESBL-E was positive for at least two consecutive time points of an infant, this infant is classified as “persistent” for this ESBL-E. If no time points were identified as ESBL-E positive, this infant is classified as “negative” for ESBL-E (never positive). Scenarios between persistent and negative are classified as “transient”, and ESBL-E typing is noted for each infant. Individual ESBL genes (e.g. EC-15, OXY-1-1) were collapsed into beta-lactamase gene family (e.g. EC family, OXY-family) because sequences of the genes within the same family have > 80% nucleotide identity (**Figure S17**). With > 0.8 as the breadth of coverage threshold, it is not distinguishable at the allele level.

Details of Data processing, statistical analyses, and visualization are described in **Supplementary Methods.**

## Data Availability

The datasets generated during and/or analyzed during the current study are available in the NCBI Sequence Reads Archive (SRA) under BioProject PRJNA1515408. Metadata that may link data back to individuals was omitted due to participant protected health. Limited, deidentified clinical and demographic data are available from authors upon request.

## Code Availability

All code for data cleaning, analysis, and visualization in this current study is publicly available at https://github.com/hartmann-lab/Long-ACQUIRE_manuscript.

## Acknowledgements

This study was funded by a Dixon Translational Research Grant and the Buffett Institute for Global Affairs Northwestern University. Additional salary support was received from National Institute of Allergy and Infectious Diseases/K23 AI139337 (L.B.M.) and K08AI123524 (M.A.). During this work, L.B.M. received salary support from NIH (K23AI139337).

This research was supported in part through the computational resources and staff contributions provided by the Genomics Compute Cluster, which is jointly supported by the Feinberg School of Medicine, the Center for Genetic Medicine, Feinberg’s Department of Biochemistry and Molecular Genetics, the Office of the Provost, the Office for Research, the Weinberg College of Arts and Sciences, and Northwestern Information Technology. The Genomics Compute Cluster is part of Quest, Northwestern University’s high-performance computing facility, with the purpose of advancing research in genomics. We also acknowledge the Rush Genomics and Microbiome Core Facility for metagenomic sequencing of the samples.

We acknowledge the support of Sebastian Otero, clinical research coordinator, the Stanley Manne Children’s Research Institute of Lurie Children’s, and Jiafeng Li for her statistical input. REDCap secure database and infrastructure were supported by Northwestern University Clinical and Translational Science Institute.

