## Supplementary Information for "Persistence of Extended Spectrum β-Lactamase-Producing Enterobacterales in the Gut Microbiome of Healthy Newborns"

### Supplementary Figures

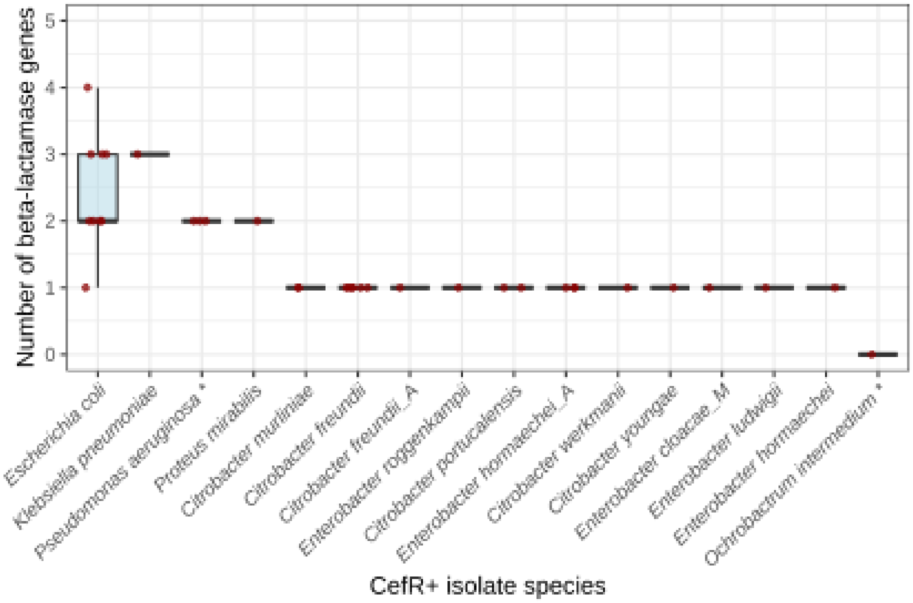


**Figure S1**. Number of β-lactamase (BL) genes carried by ceftriaxone resistant (CefR+) isolates from all samples collected from the cohort. Species labeled with “*” (*Pseudomonas aeruginosa* and *Ochrobactrum intermedium*) are not Enterobacterales.

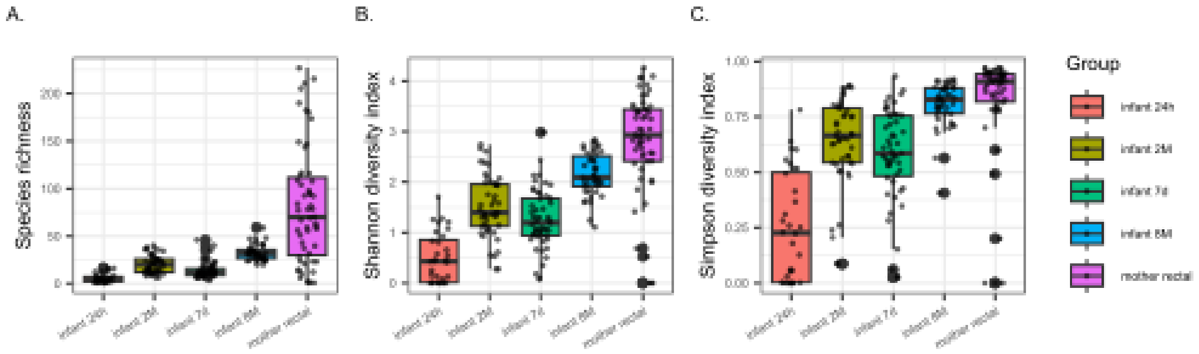


**Figure S2.** Alpha diversity indexes of mother and infant gut microbiome samples at different time points. ANOVA test $p$ values are $3.6\times10^{-28}$, $4.1\times10^{-37}$, and $1.5\times10^{-28}$ for richness, Shannon diversity index, and Simpson’s diversity index, respectively.

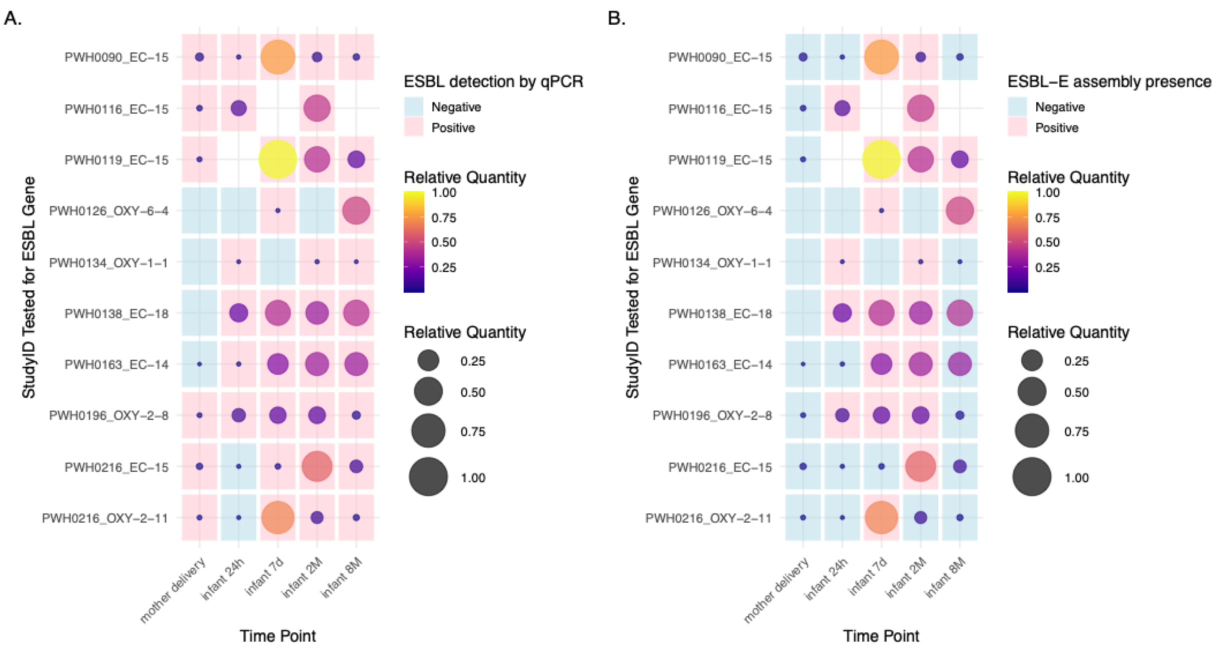


**Figure S3.** Relative quantity converted from quantitative PCR (qPCR) cycle threshold values for StudyIDs that have at least two time points classified as ESBL-E positive for assembly-based method. Samples are labeled with ESBL detection by qPCR (A) or ESBL-E assembly in the metagenome (B).

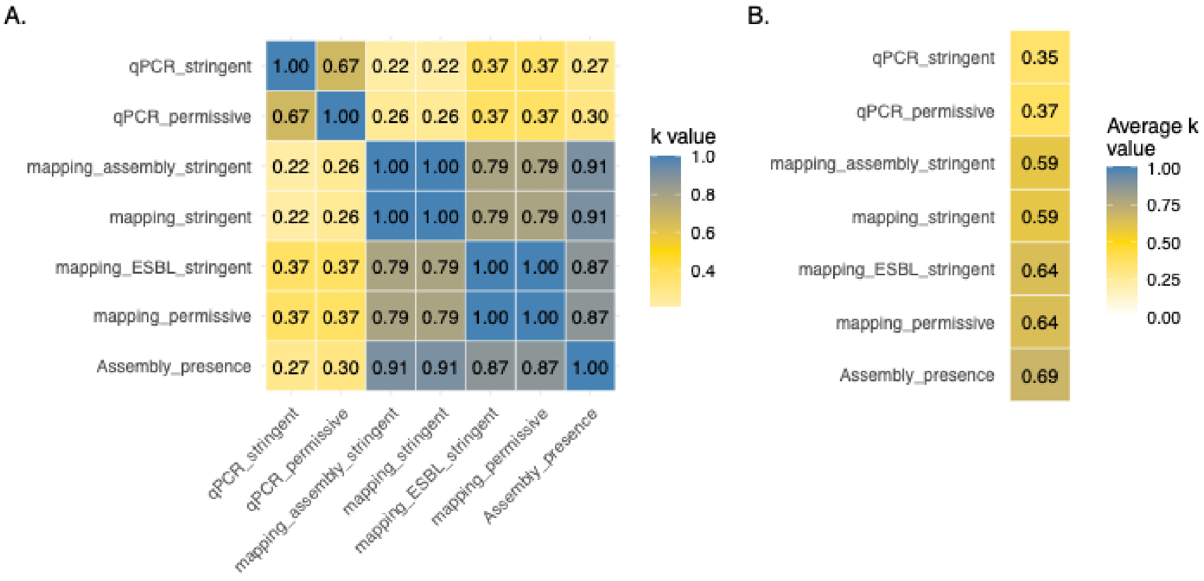


**Figure S4.** Agreements of different ESBL-E classification approaches with different parameters. Pairwise Cohen's kappa coefficient (κ value) between different methods (A) and the average κ value for each method.

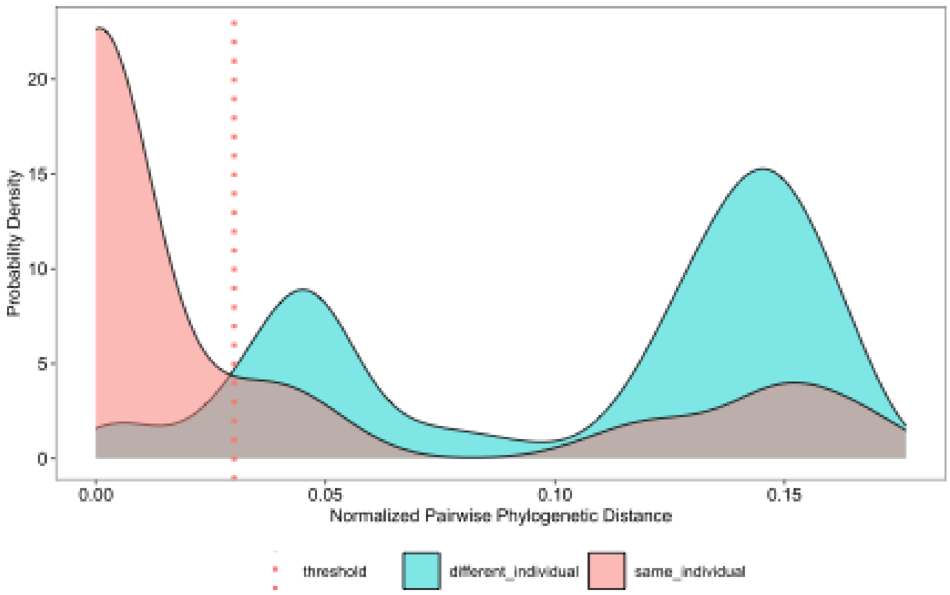


**Figure S5.** Probability density distribution of normalized pairwise phylogenetic distance of *E. colli* strains among the infants in the cohort.

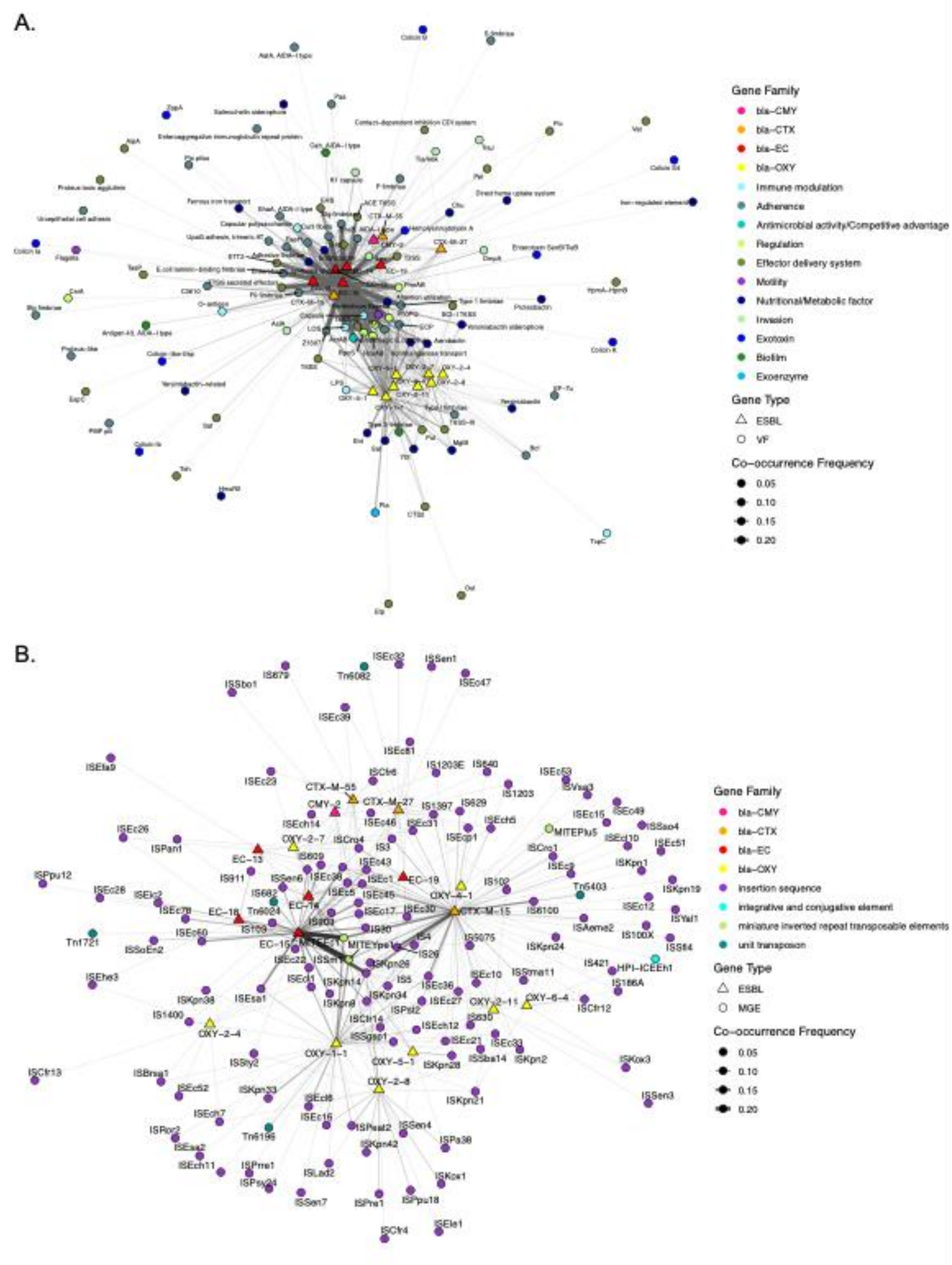


**Figure S6**. Co-occurrence network based on ESBL genes and VF genes (A) or MGE genes (B) co-localization in ESBL-E genomes (including both culture-WGS generated genomes and MAGs). Edge width and transparency represent the co-occurrence frequency of ESBL and MGE genes, defined as the proportion of genomes in which both genes are present.

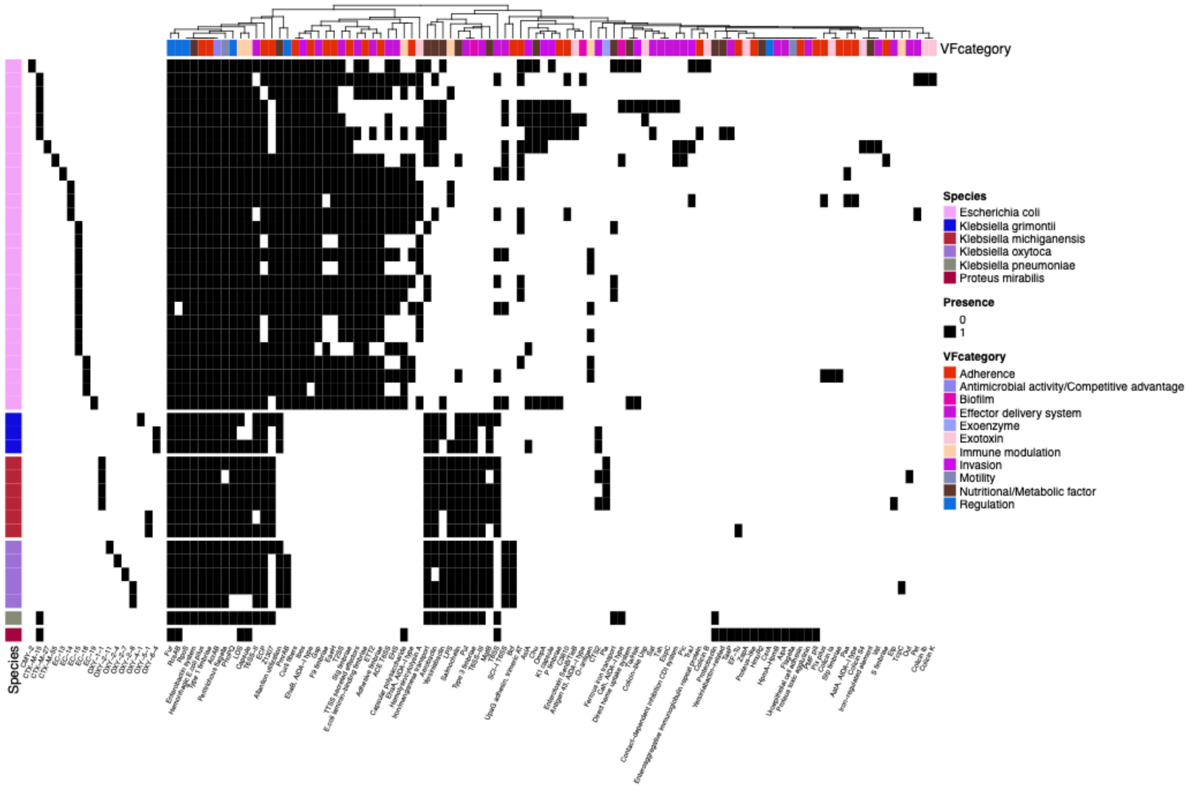


**Figure S7.** Presence/absence of VF in ESBL-E genomes. VF are clustered by their presence/absence matrix and annotated by VF category.

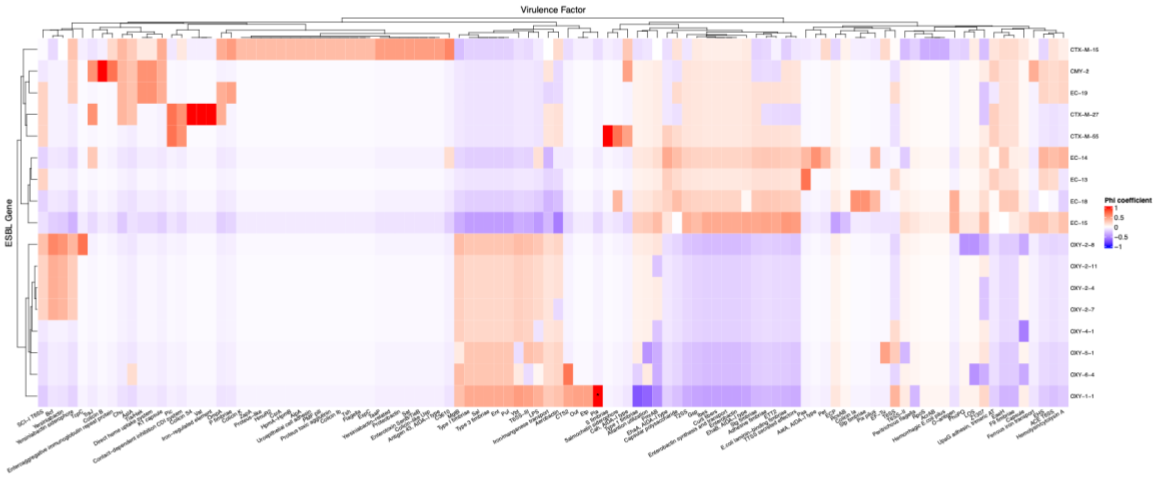


**Figure S8.** Phi coefficients between ESBL genes and VF genes in culture-WGS generated genomes and MAGs. The tile labeled with "*" represent the association with Fisher's exact test $p < 0.05$. The association between bla-OXY-1-1 and VF gene Pla has $p = 0.016$.

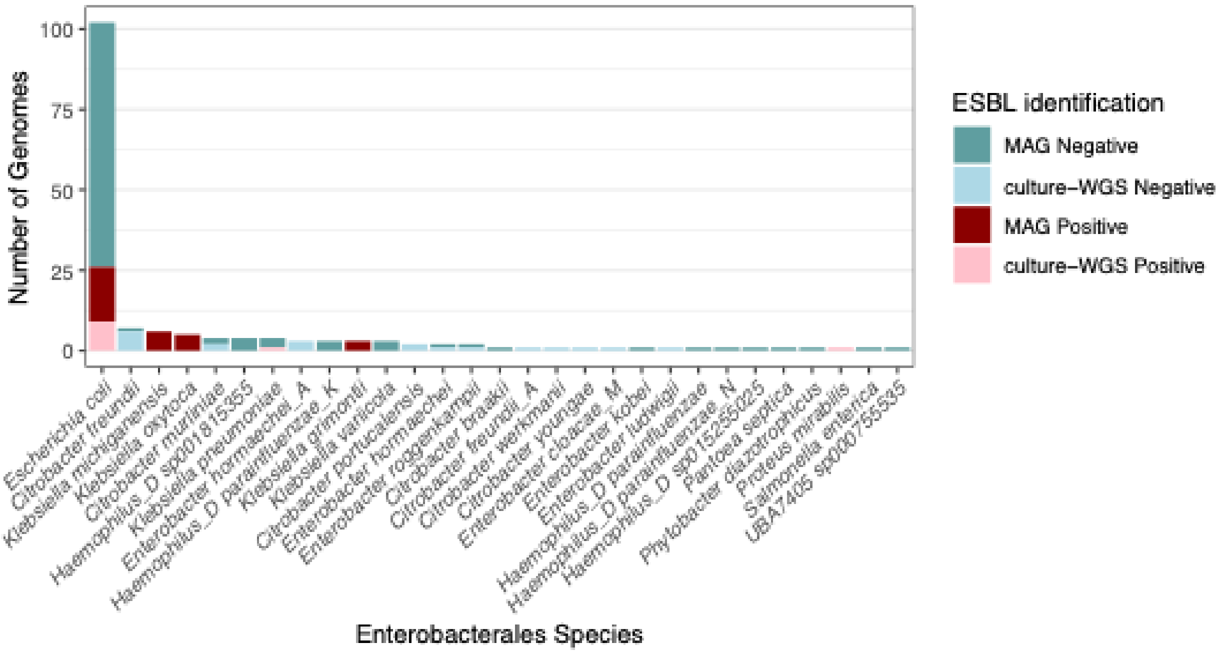


**Figure S9.** Number of Enterobacterales genomes of each species grouped by ESBL gene identification result and genome generation methods.

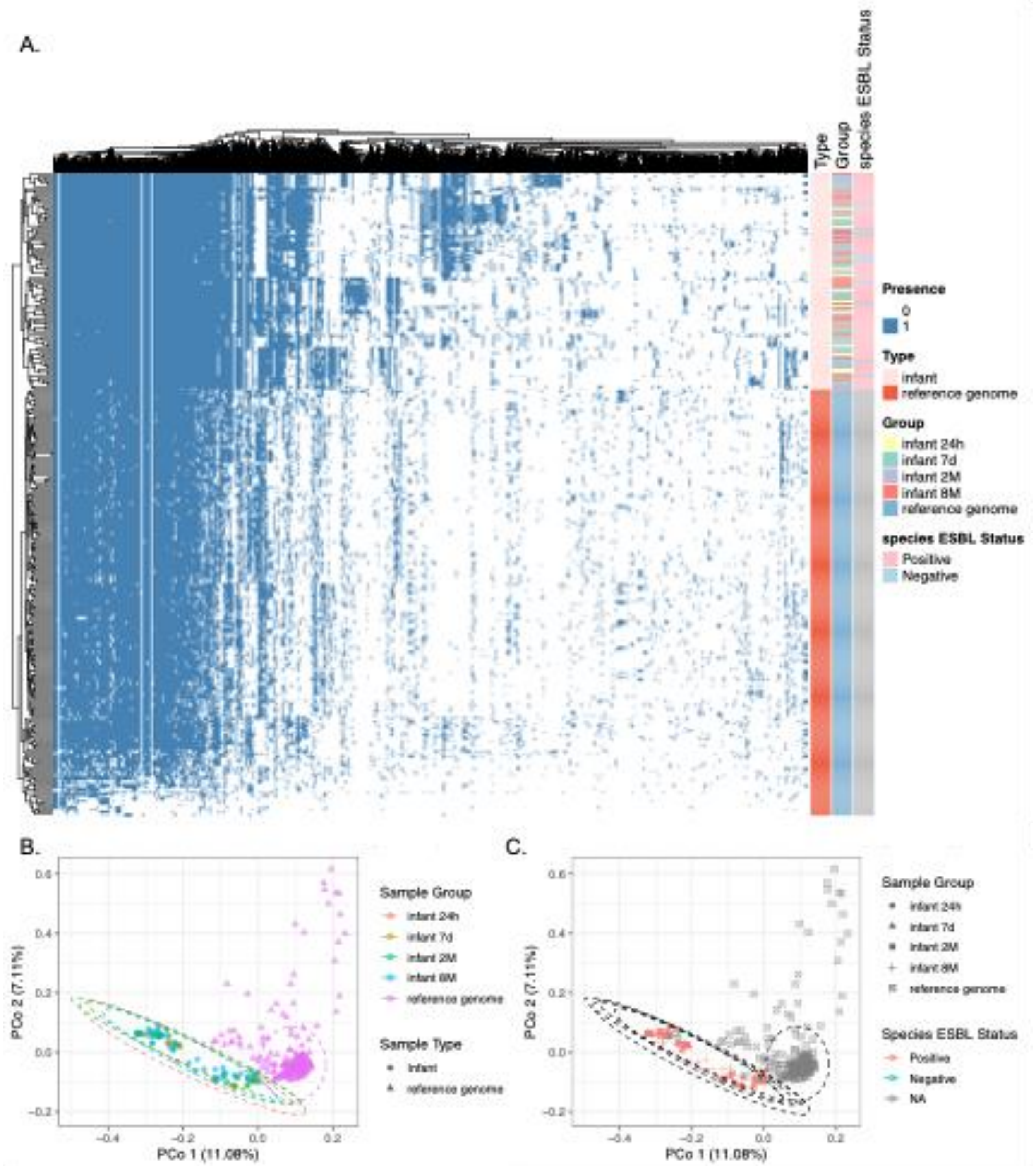


**Figure S10.** Gene composition profiling of *E. coli* strains in metagenomic samples. (A) Hierarchical clustering based on gene presence/absence matrix. PCoA based on Jaccard distance of *E. coli* gene presence/absence matrix labeled by sample group (B) and the ESBL status of *E. coli* of each sample (C).

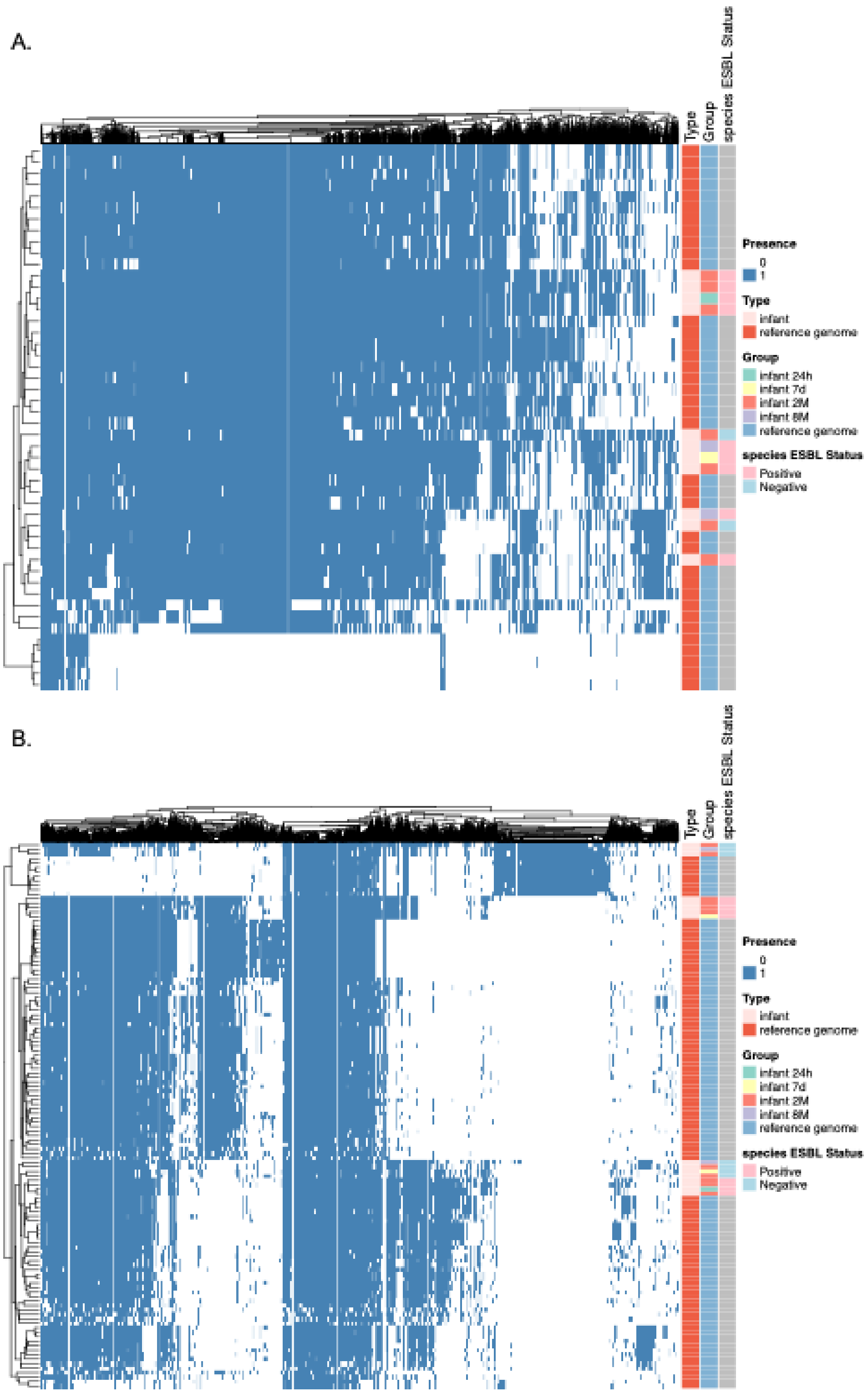


**Figure S11.** Gene composition profiling of *K. michiganensis* (A) and *K. oxytoca* (B) strains in metagenomic samples. Hierarchical clustering is based on gene presence/absence matrix.

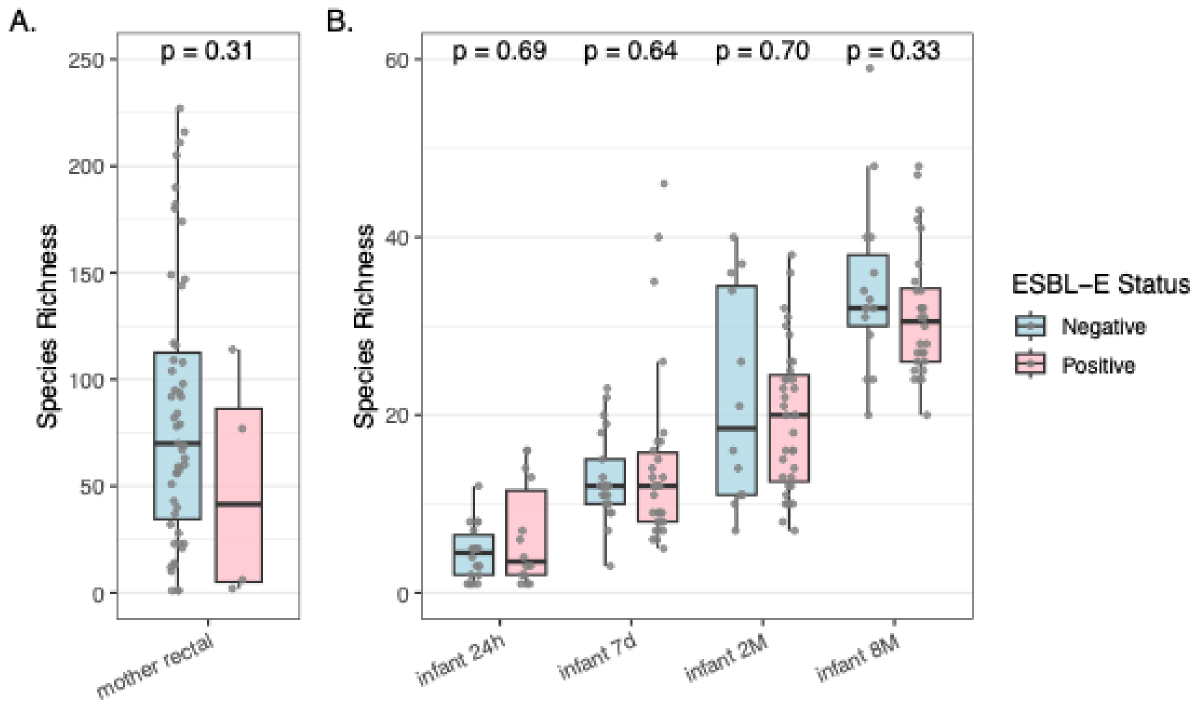


**Figure S12.** Alpha diversity (richness at species level) of ESBL-E positive versus ESBL-E negative gut microbiome for mother samples (A) and infant samples over different time points (B). Wilcoxon rank-sum test *p* values are labeled in figures.

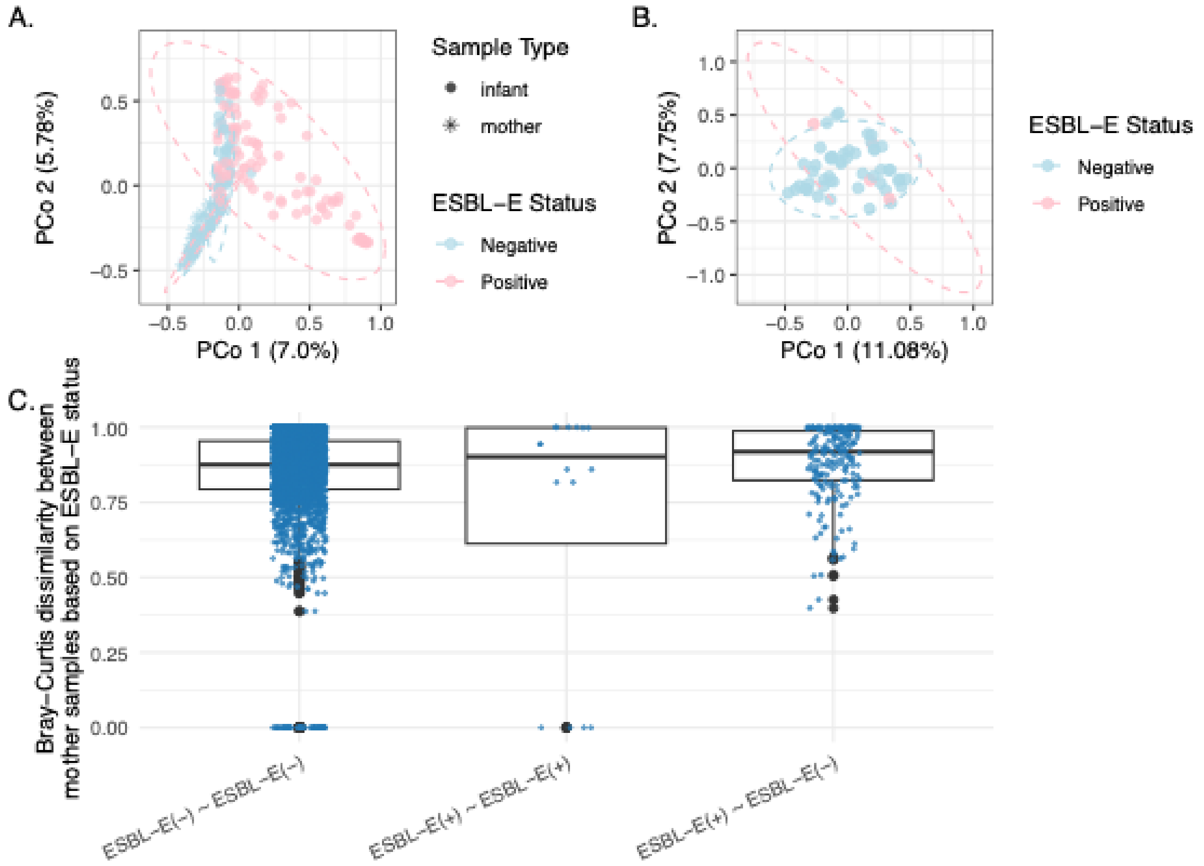


**Figure S13.** Principal coordinate analysis of gut microbial community Bray-Curtis dissimilarity for all mother and infant samples (A) and mother samples (B) labeled by ESBL-E status. PERMANOVA test permutations = 9999, $p$ = 0.66. Bray-Curtis dissimilarities among ESBL-E negative, among ESBL-E positive, and between ESBL-E positive and negative mother gut microbiome samples (C).

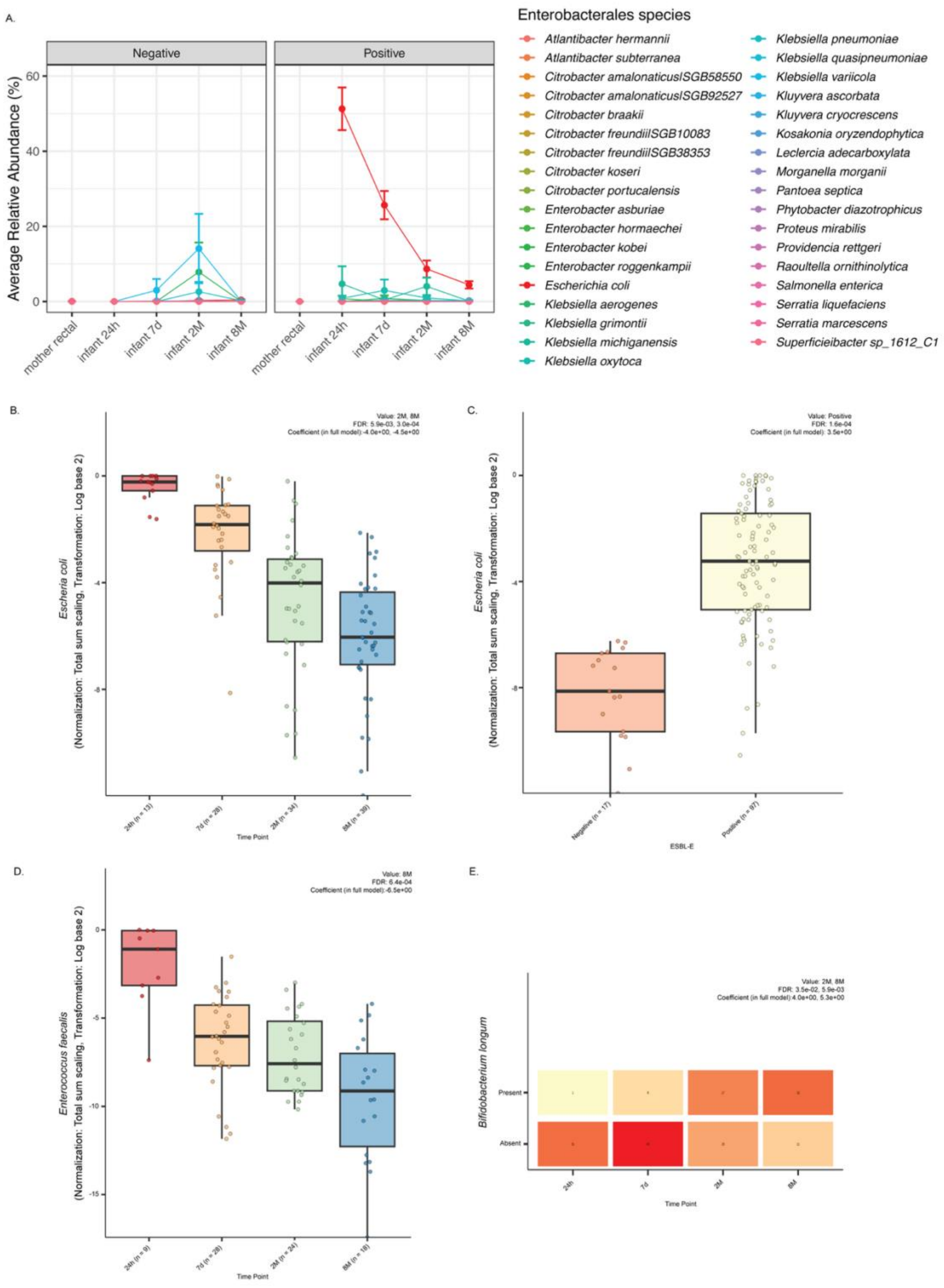


**Figure S14.** Relative abundance of Enterobacterale species in mother and infant metagenome samples over time (A). Taxa with significantly differential abundances (B, C, and D) or prevalence (E) between different time points (B, D, and E) or ESBL-E status (C) based on MaAsLin3 outputs. False discovery rate adjusted *p* values are labeled as FDR.

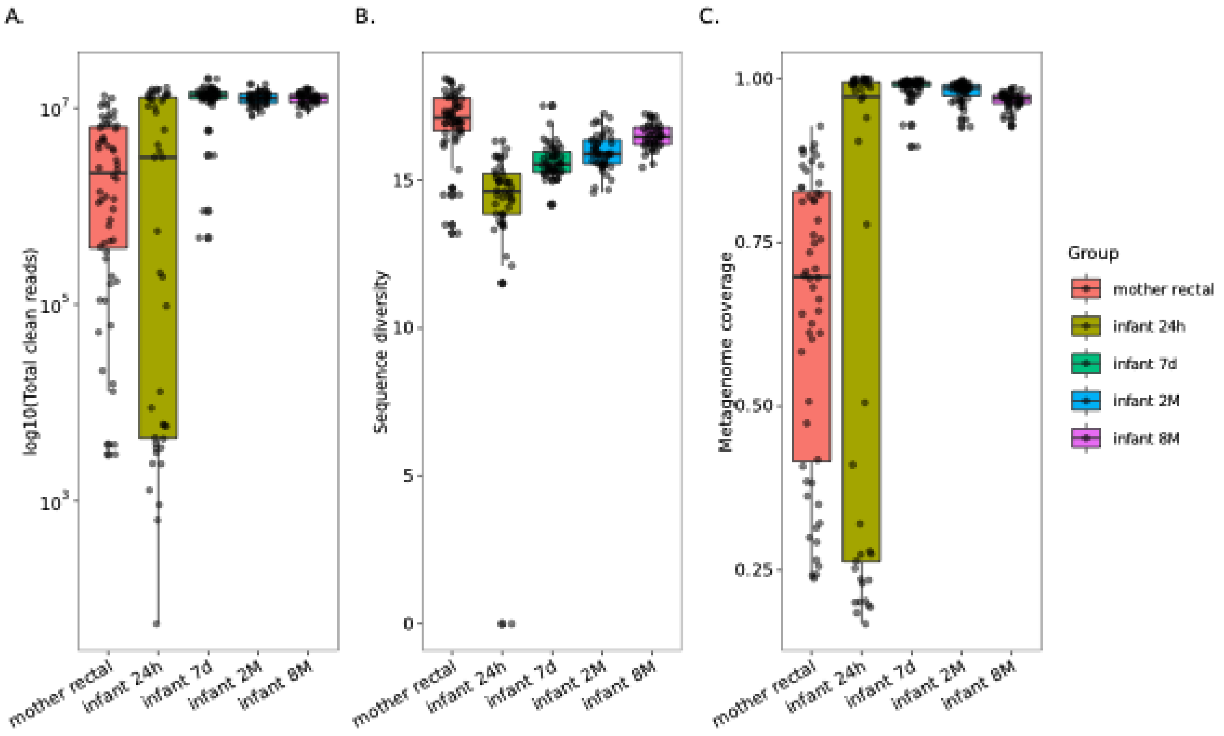


**Figure S15.** Metagenome coverage and total cleaned reads of mother and infant gut microbiome samples.

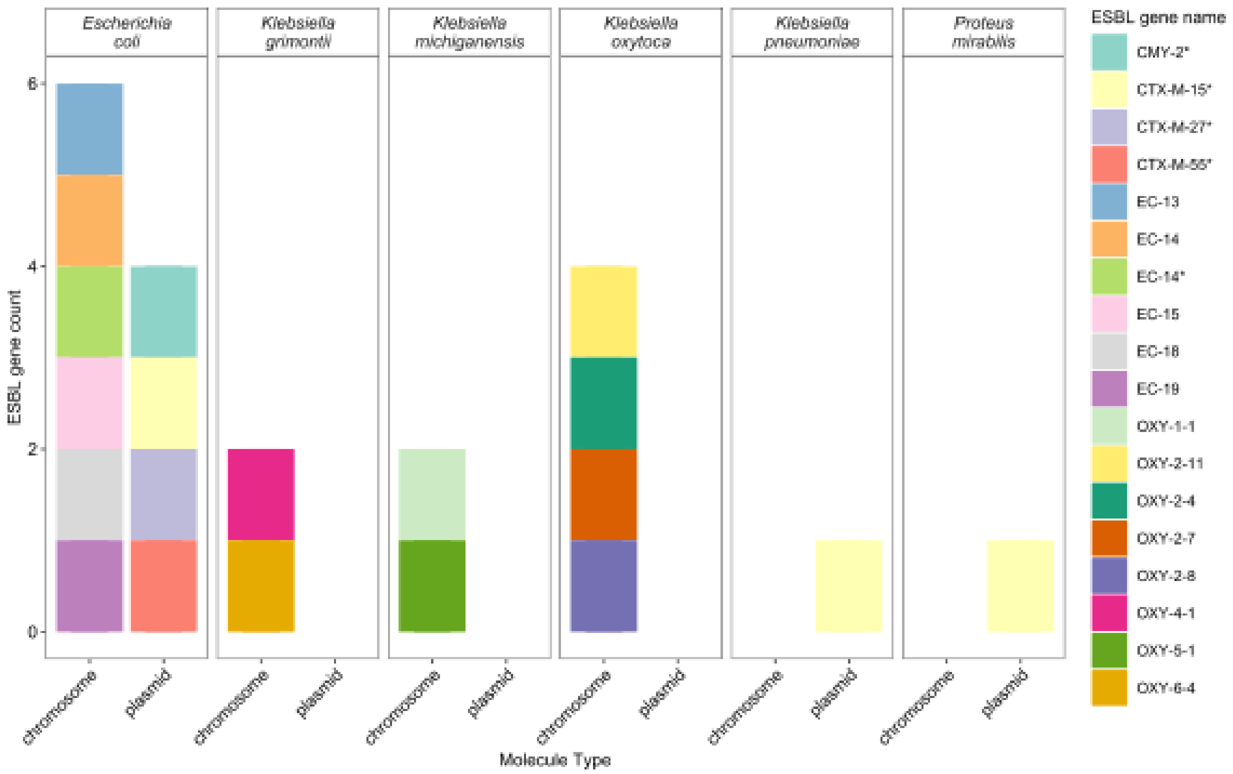


**Figure S16.** Molecule type (plasmid or chromosome) of the scaffolds that carry the ESBL genes in the Enterobacterales genomes. ESBL genes labeled with "*" are present in plasmid contigs.

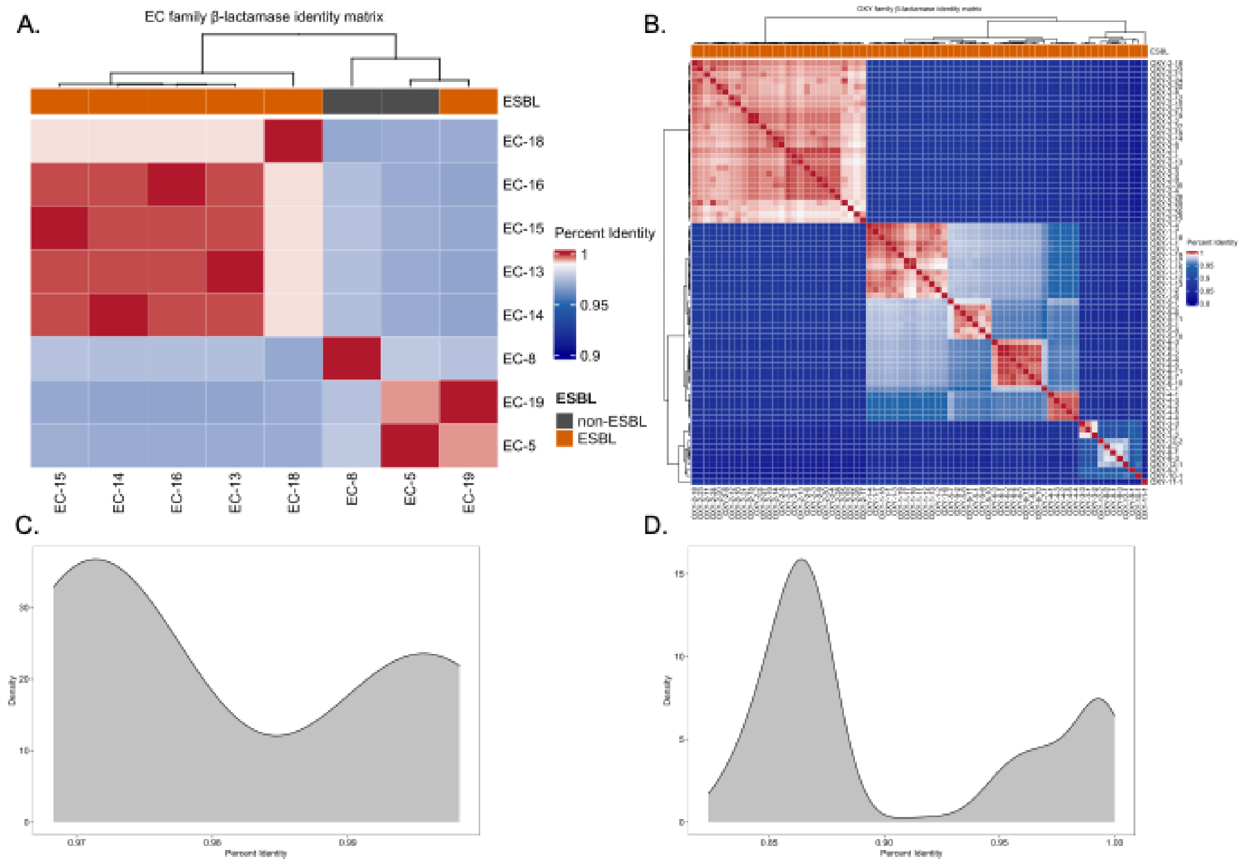


**Figure S17.** Nucleotide sequence pairwise percent identity and its distribution for ESBL genes in EC (A, C) and OXY (B, D) beta-lactamase gene families.

### Supplementary Tables

**Table S1.** Persistence status of individual infants.

Supplementary_Table_S1_Persistence_Status_per_StudyID.xlsx

**Table S2.** Pairwise Wilcox test *p* values for Bray-Curtis dissimilarities between ESBL-E positive and negative infant gut microbiome for different time points.

| | Infant 24h | Infant 7d | Infant 2M |
|---|---|---|---|
| Infant 7d | $< 2\times10^{-16}$ | | |
| Infant 2M | $< 2\times10^{-16}$ | $2.9\times10^{-6}$ | |
| Infant 8M | $< 2\times10^{-16}$ | $< 2\times10^{-16}$ | $< 2\times10^{-16}$ |

**Table S3.** Microbiome Multivariable Associations with Linear Models statistics of pathways in the infant gut microbiome metagenome.

Supplementary_Table_S3_MaAsLin3_Pathways.xlsx

### Supplementary Methods

#### *Ceftriaxone resistant isolates culturing and selection*

Parental vaginal and rectal swabs (BD BBL CultureSwab EZ sterile polyurethane, single swab) were collected during labor after consent was taken and were immediately immersed in Eppendorf tubes containing 500 µL of Luria broth (LB), 50 µg/mL of ampicillin, and 8 µg/mL of vancomycin. A pea-sized amount of stool was collected from soiled infant diapers and immersed in the culture tubes mentioned above. All LB samples were incubated overnight for 14–16 hours at 37°C in a shaking incubator and then plated on MacConkey agar with 50 µg/mL of ampicillin (growth indicating AmpR-E), as well as MacConkey agar with ceftriaxone mixed into it at a concentration of 20 µg/mL (indicating CefR-E). Plates were incubated overnight at 37°C, and growth of colonies on the plates was assessed.

#### *Whole genome sequencing sample processing*

CefR+ isolates went through whole genome sequencing. For both parental and infant stool samples that grew in the presence of ceftriaxone, single bacterial colony isolates were sequenced. Genomic DNA was extracted using the Wizard Genomic DNA Purification Kit (Promega). The isolated DNA was subjected to sequencing library preparation. For this step, 1 ng of DNA was used to start the process using the Illumina Nextera XT DNA Library Preparation Kit. DNA tagmentation, bead-based cleanup, polymerase chain reaction (PCR) amplification of tagmented DNA, PCR purification, and library quantification, quality check, and normalization were done. The Illumina MiSeq NGS System was used to sequence the libraries on a version 3 flow cell to produce paired-end 300-bp reads They were processed through genome assembly pipeline on the web server of Bacterial and Viral Bioinformatics Resource Center (https://www.bv-brc.org/) using the default settings.

#### *Shotgun metagenomic sequencing sample processing*

DNA samples of mother and infant gut microbiome were sequenced using [platform] with 150 base paired end reads. Resulting raw sequences went through quality control using Kneaddata v0.12.0 (https://huttenhower.sph.harvard.edu/kneaddata) with adaptor and host sequence removal (human reference database version hg37dec_v0.1). BBMap[1] v39.01 was used to repair unpaired fastq files with clean reads from Kneaddata for downstream analyses. Metagenome diversity and coverage were estimated using Nonpareil[2,3] v3.5.5. Taxonomy profiling of mother and infant gut microbiome was performed using MetaPhlAn[4] v4.1.0 with database mpa_vJun23_CHOCOPhlAnSGB_202403.

Metagenome-assembled scaffolds were obtained using SPAdes[5] v3.15.3 with "--meta" mode[6]. MetaWRAP[7] v1.3.2 pipeline was used to generate metagenome-assembled genomes (MAG) from the scaffolds. Initial binning was conducted with "--maxbin2 --concoct --metabat2" methods. The resulting bins were then refined with completeness > 80% and contamination < 10% and reassembled into final MAGs.

***Assembly taxonomic classification and functional annotation***

For WGS generated genomes, MAGs, and SPAdes assembled scaffolds, GTDB-kt47 v2.1.1 was used to assign taxonomy classification for Enterobacterales assembly identification. Resistance Gene Identifier (RGI) v6.0.3 "main" mode was used to identify AMR genes from the assemblies with the Comprehensive Antibiotic Resistance Database (CARD)48 v4.0.1 and filtered with best identities > 90% and coverage of reference gene length > 95%.

***Real-time quantitative polymerase chain reaction (qPCR) experiments***

Real-time quantitative PCR was performed using ESBL gene primers EC-14, EC-15, EC-18, OXY-6-4, OXY-1-1, OXY-2-11, and OXY-2-8 (Integrated DNA Technologies) as listed in **Table S4**. Each reaction contained 10μl 2x SsoAdvanced Universal SYBR Green Supermix (Bio-Rad), 0.7μl of each primer (final concentration 350 nM), 1μl template DNA, and 7.6μl Cytiva HyPure Molecular Biology Grade Water (Fisher Scientific) for a total reaction volume of 20μl. A QuantStudio 6 Flex real-time PCR system was used for thermal cycling with an activation and DNA denaturation step of 50.0 °C for 2 min and 98.0 °C for 3 min, 40 cycles of 98.0 °C for 15 sec and 55.0 °C for 1 min, and a Melt Curve Analysis step of 95.0 °C for 15 sec, 65.0 °C for 1 min, and 95.0 °C for 15 sec.

**Table S4**. ESBL gene primers used in qPCR experiments

| ESBL-E ID | Forward (5' to 3') | Reverse (5' to 3') |
|---|---|---|
| *Escherichia coli* EC-14 | GTCTTCCAGCCACTCAAACT | GCATCACTATACCCAGCTCTTT |
| *Escherichia coli* EC-15 | GTCTTCCAGCCACTCAAACT | GCATCACTATACCCAGCTCTTT |
| *Escherichia coli* EC-18 | AGGGATACAACTGGCACAATC | ACGCGACATAGCTACCAAATC |
| *Klebsiella grimontii* OXY-6-4 | CCATCCCGATGTGGTGAATAA | GTTGGTGGTGCCGTAATCT |
| *Klebsiella michiganensis* OXY-1-1 | GCAATCCAGAGGTGGTGAATA | GTTGGTGGTGCCGTAATCT |
| *Klebsiella oxytoca* OXY-2-11 | CGACAATACGGCGATGAATCT | TCATTGGTGGTGCCGTAATC |
| *Klebsiella oxytoca* OXY-2-8 | CGACAATACGGCGATGAATCT | TCATTGGTGGTGCCGTAATC |

***Comparisons of different ESBL-E classification approaches with different parameters***

The cycle threshold (CT) values for the qPCR experiment targeting different ESBL genes were converted to relative quantities (RQ) for qualitative ESBL genes detection in the gut microbiome samples. The formula for the conversion is:

$$RQ = \sqrt{\frac{2^{-CT}}{\max(2^{-CT})}}$$

For qPCR-based ESBL detection, permissive and stringent criteria were applied. Permissive detection means that ESBL is classified as positive when sample average CT values are lower than the average control sample CT values or lower than 40. Stringent detection means that ESBL is classified as positive when sample average CT values are lower than 35.

For ESBL-E assembly presence-based ESBL-E status classification, a sample is classified as ESBL-E positive for this ESBL-E typing (for example, *E.coli* assembly carrying EC-15 ESBL gene) if an ESBL-E assembly has resulted from this metagenome sample.

For ESBL-E assembly mapping-based ESBL-E status classification, permissive and stringent criteria were applied to both ESBL-E assembly detection and ESBL gene detection. The rationale for considering both ESBL-E asssembly and ESBL gene mapping was that the coverage of the specific ESBL gene within the ESBL-E assembly (a MAG or a scaffold) may not be reflected by the coverage and depth of the entire ESBL-E assembly mapping result. **Figure S18** depicts the coverage of the assembly and of the ESBL gene. To make sure that our results are specific for ESBL genes carried by the Enterobacterales assemblies, sufficient coverage fraction of ESBL genes is necessary. The following table shows the parameters applied for mapping-based ESBL-E classification.

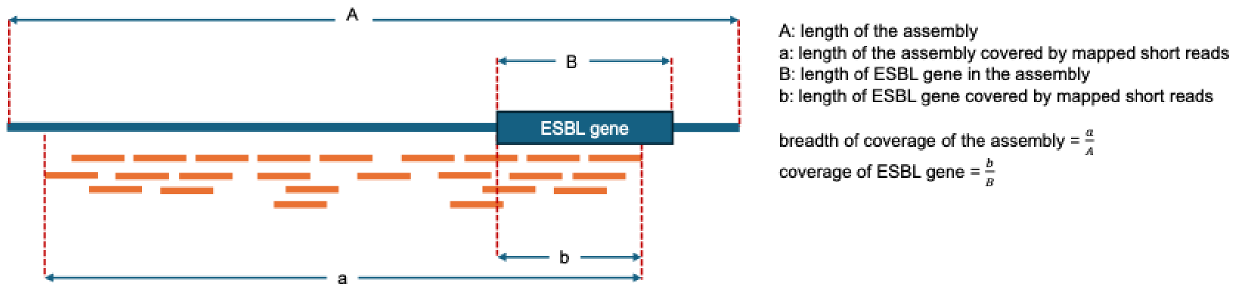


**Figure S18.** Diagram showing the definition of breadth of coverage of the assembly and coverage of ESBL gene in the assembly.

**Table S5.** Thresholds used for different criteria of ESBL-E assembly mapping-based classification methods.

| Criteria | ESBL-E assembly mapping | | ESBL gene mapping |
|---|---|---|---|
| | Truncated average depth normalized to per million clean reads | Breadth of coverage (0-1) | Coverage fraction (0-1) |
| Permissive | > 0.2 | > 0.25 | > 0.5 |
| Assembly stringent | > 0.5 | > 0.5 | > 0.5 |
| ESBL stringent | > 0.2 | > 0.25 | > 0.8 |
| Stringent | > 0.5 | > 0.5 | > 0.8 |

The degree of agreement between different classification approaches with different criteria was estimated using pairwise Cohen's kappa coefficient (κ value). Since qPCR was performed only for 9 StudyIDs where at least 2 infant time points resulted in ESBL-E assemblies, κ values were only calculated for these samples across different approaches. For mapping-based ESBL-E status classification, we would prioritize the criteria that both have a better agreement with the experimental-based (qPCR) method and are specific enough for ESBL gene detection. Therefore, "ESBL stringent" criteria were chosen for the ESBL-E assembly mapping-based classification approach.

***Data processing, statistical analyses, and visualization***

Clinical and demographic data comparisons were performed using the Wilcoxon rank-sum test or Kruskal–Wallis test, as appropriate. For categorical variables, Fisher's exact test or Chi-square tests were applied. Analysis of variance (ANOVA), principal coordinates analysis (PCoA) on Bray-Curtis dissimilarity matrices, permutational multivariate analysis of variance (PERMANOVA), Kruskal-Wallis rank sum test, Wilcoxon rank sum test, microbiome multivariable associations with linear models (MaAsLin3, R package maaslin3 v0.99.7)[59], and Fisher's exact test for phi coefficient (co-occurrence associations for presence/absence of ESBL genes, MGEs, and VFs in Enterobacterales genomes) were performed in R 4.4.0[60]. StrainPhlAn4[43,61] was performed on infant gut microbiome metagenomic sequencing samples for strain-level resolution of selected Enterobacterales species. Hierarchical all-against-all association (HAllA)[62] was performed on abundance tables of species-level genome bins and ESBL-E assemblies. Visualizations were created in R with ggplot2[63] v4.4.0, ggtree[64] v3.8.2, ComplexHeatmap[65] v2.22.0, ggalluvial[66,67] v0.12.6, maaslin3[59] v0.99.7, and patchwork[17] v1.3.2.
